# Shared and unique chromatin accessibility and 3D functional interactions of substance use disorder loci implicate cell types beyond the brain

**DOI:** 10.1101/2024.07.18.24310649

**Authors:** Khanh B. Trang, Alessandra Chesi, Sylvanus Toikumo, James A. Pippin, Matthew C. Pahl, Joan M. O’Brien, Laufey T. Amundadottir, Kevin M. Brown, Wenli Yang, Jaclyn Welles, Dominic Santoleri, Paul M. Titchenell, Patrick Seale, Babette S. Zemel, Yadav Wagley, Kurt D. Hankenson, Klaus H. Kaestner, Stewart A. Anderson, Matthew S. Kayser, Andrew D. Wells, Henry R. Kranzler, Rachel L. Kember, Struan F.A. Grant

**Affiliations:** Center for Spatial and Functional Genomics, Children’s Hospital of Philadelphia, Philadelphia, PA, USA; Division of Human Genetics, Children’s Hospital of Philadelphia, Philadelphia, PA, USA; Department of Pathology and Laboratory Medicine, Perelman School of Medicine, University of Pennsylvania, Philadelphia, PA, USA; Mental Illness Research, Education and Clinical Center, Crescenz Veterans Affairs Medical Center, Philadelphia, PA, USA; Department of Psychiatry, Perelman School of Medicine, University of Pennsylvania, Philadelphia, PA, USA; Division of Gastroenterology, Hepatology, and Nutrition, Children’s Hospital of Philadelphia, PA, USA; Scheie Eye Institute, Department of Ophthalmology, Perelman School of Medicine, University of Pennsylvania, Philadelphia, Pennsylvania, PA, USA; Penn Medicine Center for Ophthalmic Genetics in Complex Disease, Perelman School of Medicine, University of Pennsylvania, Philadelphia, Pennsylvania, PA, USA; Laboratory of Translational Genomics, Division of Cancer Epidemiology and Genetics, National Cancer Institute, Bethesda, MD, USA; Department of Medicine, Division of Translational Medicine and Human Genetics, Perelman School of Medicine, University of Pennsylvania, Philadelphia, PA, USA; Institute for Diabetes, Obesity and Metabolism, Perelman School of Medicine, University of Pennsylvania, Philadelphia, PA, USA; Department of Cell and Developmental Biology, Perelman School of Medicine, University of Pennsylvania, Philadelphia, PA, USA; Department of Physiology, Perelman School of Medicine, University of Pennsylvania, Philadelphia, PA, USA; Department of Pediatrics, Perelman School of Medicine, University of Pennsylvania, Philadelphia, PA, USA; Department of Orthopedic Surgery, University of Michigan Medical School Ann Arbor, MI, USA; Department of Genetics, Perelman School of Medicine, University of Pennsylvania, Philadelphia, PA, USA; Department of Child and Adolescent Psychiatry, Children’s Hospital of Philadelphia, Philadelphia, PA, USA; Department of Neuroscience, Perelman School of Medicine, University of Pennsylvania, Philadelphia, PA 19104, USA; Chronobiology Sleep Institute, Perelman School of Medicine, University of Pennsylvania, Philadelphia, PA 19104, USA; Institute for Immunology, Perelman School of Medicine, University of Pennsylvania, Philadelphia, PA 19104, USA; Division of Endocrinology and Diabetes, The Children’s Hospital of Philadelphia, Philadelphia, PA, USA

**Keywords:** Substance Use Disorders, Genome-Wide Association Study, Type 2 Diabetes Mellitus, Mendelian Randomization Analysis, Epigenomics

## Abstract

**Objective:** Recent genome-wide association studies (GWAS) have revealed multiple loci for substance use disorders (SUDs), including evidence that the traits overlap in genetic etiology. However, the extent of underlying SUD causal variants, effector genes, and cellular contexts, remains unclear. Recent clinical trials of glucagon-like peptide-1 (GLP-1) receptor agonists have shown promising effects on alcohol and tobacco consumption, raising the question of whether SUD genetic risk resides in metabolic as well as neural cell types. We sought to map the cell-type-specific genetic architecture of four SUDs and to test whether the observed metabolic enrichment reflects shared causal biology with type 2 diabetes (T2D).

**Methods:** We integrated our 3D genomic datasets (high-resolution promoter-focused Capture-C/Hi-C, ATAC-seq, RNA-seq) from 59 diverse human cell types with recent GWAS summary statistics for alcohol (AUD), tobacco (TUD), opioid (OUD), and cannabis use disorder (CanUD), using Stratified LD regression (S-LDSC). We performed local genetic correlation analysis (LAVA) between SUDs and T2D, multivariate integration (DIABLO) of gene expression and chromatin accessibility at shared loci, and subsequent bidirectional two-sample Mendelian randomization (MR) to test for causal relationships between T2D and each SUD.

**Results:** After per-trait false discovery rate (FDR) correction, TUD showed significant heritability enrichment in pancreatic *β*-cells (4.6-fold), α-cells (4.0-fold), hypothalamic neurons, and iPSC-derived neurons. CanUD showed FDR-significant enrichment concentrated in hypothalamic neural progenitor cells (7.4-fold). LAVA identified 1,550 loci with significant local genetic correlations, 114 specifically between T2D and at least one SUD. Subsequent MR revealed that genetic liability for T2D was protective against AUD (inverse-variance weighted *β* =-0.027, *P*-values = 1.6×10^-5^, 463 instruments), consistent across four of five MR methods and free of directional pleiotropy. Cell-cell interaction modeling implicated distinct neuro-endocrine signaling axes disrupted by each SUD.

**Conclusions:** These findings implicate a pancreatic-SUD axis, validated by orthogonal MR evidence, that reframe SUDs as multi-system disorders, and provides a genetic-mechanistic rationale for GLP-1-based therapeutic strategies targeting addiction.

**Significant Outcomes:**

1. Local genetic correlation analysis identifies 114 genomic loci shared between T2D and SUDs; multivariate integration at these loci reveals conserved regulatory architecture between neural and pancreatic cells operating through distinct neuro-endocrine signaling pathways.
2. Tobacco use disorder heritability is significantly enriched (FDR < 0.05) in pancreatic α-and β-cells alongside neural lineages, establishing a multi-system genetic architecture that extends beyond the brain.
3. Bidirectional Mendelian randomization demonstrates that genetic liability for T2D is causally protective against alcohol use disorder (*P*-values = 1.6 × 10^-5^), providing orthogonal validation of the pancreatic-SUD axis identified by chromatin-based enrichment.

**Limitations:**

1. Analyses were restricted to European-ancestry GWAS; generalizability to other populations remains untested.
2. Ligand-receptor signaling inferences (CellChat) are computational predictions from curated databases, not experimentally validated interactions; directional claims require future *in vivo* confirmation.
3. Drug-target (cis) MR at the *GLP1R* locus did not reach significance for any SUD, indicating that the broader pancreatic axis – rather than GLP-1 receptor signaling alone – likely mediates the observed genetic overlap.

## INTRODUCTION

Substance use disorders (SUDs) are associated with many adverse outcomes, representing a global health crisis. SUD severity varies from mild to severe, and adverse outcomes generally correlate with severity. Alcohol, socially accepted and widely available, is one of the most misused substances worldwide(^1^), leading to significant health, social, and economic costs. Excessive alcohol consumption is a major cause of premature death and disability, particularly among individuals aged 20-39, with approximately 13.5% of deaths in this group linked to alcohol. Alcohol use disorder (AUD) affects 10.5% of the US population over age 12, contributing to over 178,000 deaths annually(^2, 3^). Tobacco use disorder (TUD), the most prevalent SUD, affects 22.3% of the global population and causes nearly 8 million preventable deaths annually(^4^). Opioid use disorder (OUD) prevalence has surged due to prescription misuse and illicit opioids like fentanyl, resulting in approximately 81,806 US overdose deaths in 2022(^5^). The prevalence of cannabis use disorder (CanUD) has also risen with increasing legalization and is associated with morbidity, cognitive impairments, and schizophrenia(^6–9^).

SUDs show strong familial inheritance patterns(^10^), with twin studies estimating an average heritability of ∼50%. AUD heritability estimates range from 0.50-0.64(^11, 12^), TUD heritability ranges from 0.30 to 0.70(^13, 14^), while CanUD heritability ranges from 0.5-0.59(^15, 16^). OUD heritability is ∼50%, with 38% of variation attributed to opioid-specific genetic factors(^17^). Beyond specific disorders(^18, 19^), broader heritable factors influence the general susceptibility to SUDs(^20^). Over the past decade, GWAS have identified numerous significant loci for SUDs(^21, 22^). The first signals associated with AUD reside near *ADH1B* and *ALDH2*(^23–31^), which encode metabolic enzymes, and has expanded to include *DRD2, GCKR, KLB* and *SLC39A8*(^25, 27, 29, 30, 32, 33^). TUD GWAS consistently link nicotine dependence to cholinergic nicotinic receptor genes, specifically at the *CHRNA5-CHRNA3-CHRNB4* locus (^22^) and *DNMT3B* loci. Recent multi-ancestral meta-analyses revealed 72 independent risk loci for TUD(^34^), and more than a thousand loci associated with various smoking phenotypes(^35^). While CanUD genetic research has been limited by smaller samples, recent meta-GWAS(^36^) revealed several loci unique both to individual ancestries, and confirmed loci near *CHRNA2*(^37^) and *FOXP2*(^38^). GWAS for OUD have implicated the *CNIH3*, *KCNG2*, *APBB2*, *RGMA*, *KCNC1*, and *OPRM1* genes(^32, 39–41^). Other large-scale analyses using multi-trait methods(^42^) have identified additional novel OUD loci(^43^).

Despite the growing number of common genetic variants identified for these SUDs, there is lack of cell-type resolution in GWAS(^44^). Furthermore, accurately interpreting these loci requires robust variant-to-gene mapping, as relying merely on linear genomic proximity often implicates the wrong causal gene and overlooks the profound cell-type specificity of regulatory interactions. standard tissue enrichment analyses (e.g., broad LDSC-SEG) have consistently emphasized the brain with minimal evidence for peripheral involvement (^45^).

The recent clinical emergence of glucagon-like peptide-1 (GLP-1) receptor agonists as potential therapeutics for AUD and TUD (^46–48^), with effect sizes comparable to first-line addiction pharmacotherapies, thus raises a fundamental question: does SUD genetic risk reside in the metabolic cell types that produce incretin hormones? If so, the pancreatic-neural axis targeted by semaglutide and tirzepatide may not be merely a pharmacological convenience but a reflection of shared causal biology.

To address this, we integrated genome-wide heritability partitioning (S-LDSC)(^49^) with cell-type-specific 3D chromatin architecture across 59 human cell types, local genetic correlation analysis (LAVA), cell-cell signaling inference, and subsequent bidirectional Mendelian randomization between T2D and SUDs. This multi-layered approach sheds light on the cellular contexts, shared regulatory architectures, and causal relationships underlying SUD genetic risk.

## METHODS AND MATERIALS

### Data and resource

**Table S1** lists the multi-omic datasets spanning immune, metabolic, neuronal, and pluripotent lineages used in prior studies from our laboratory. European-ancestry GWAS summary statistics for AUD, TUD, CanUD, and OUD, and all other traits explored in this study, are listed in **Table S2**. GWAS summary statistics were standardized using *-- merge-alleles* with HapMap3 variants. Baseline-LD model (v2.2) LD scores, plink files, allele frequencies, and regression weights for the European 1000 Genomes Phase 3 reference in GRCh38 were obtained from the Alkes Group repository.

### Multi-Omic Data Preprocessing and Functional Annotation

We integrated ATAC-seq, RNA-seq, and 3D-genomic data (Hi-C, Capture-C) across 59 cell types, using standardized processing pipelines(^50^).

Open chromatin regions (OCRs) were called using the nf-core ATAC-seq Nextflow pipeline(^51^) with Chromap alignment (^52^) and MACS2 peak calling. For single-cell ATAC-seq pancreatic data (50), fragments from the same cell identity groups were pooled per cell type and processed identically.

RNA-seq reads were aligned using STAR (^52^) with GENCODE annotation; read counts were calculated with htseq-count, transformed to TPM, and internally normalized.

Promoter Capture-C reads were processed with HICUP (^53^) and significant interactions called using CHICAGO (^54^) at 1-fragment and 4-fragment resolution (score > 5), then merged. Hi-C reads were processed with HICUP and pairtools (^55^), converted to.cool matrices via cooler (^56^), and ICE-normalized (^57^). Significant interactions were called using both Mustache(^58^) (*P*-values < 0.1) and Fit-Hi-C2(^59^) (FDR < 1×10^-6^) at 1, 2, and 4 kb resolutions; the consensus loop set was generated by merging interactions identified by both methods across resolutions.

### Functional annotation categories for S-LDSC

We categorized the genome from each cell type into six annotation categories: (i) **Total OCRs** for global assessment; (ii) **Promoter OCRs** testing direct promoter effects; (iii) **cREs** (cis-regulatory elements linked to promoters via chromatin loops) and (iv) **cREs ± 500 bp** to capture variants near regulatory element boundaries(^49^); (v) **not-cREs/Prom OCRs** as a negative control (accessible but non-looping, non-promoter regions); and (vi) **SEGs** (specifically expressed gene sets, top 10% from DESeq2(^60^) with apeglm(^61^) shrinkage, plus 100 kb flanking) as a positive control(^45^). All annotation-specific LD scores were analyzed jointly with the 63 categories of the baseline-LD model (v2.2), which controls for annotation overlap. Two statistics assess how annotations capture causal variation: partitioned heritability enrichment (with associated *P*-value) and standardized effect size *τ**\**** (^62^), which enables cross-cell-type comparisons independent of annotation size.

### Heritability and Genetic Correlation Analyses

Stratified LD Score Regression (S-LDSC) (^49^) partitioned SNP-heritability across regulatory landscapes, including promoters and distal cREs, while controlling for the baseline-LD model. Global and cell-type-specific genetic correlations (r_g_) between SUDs and metabolic traits were estimated using LDSC(^63^). Local genetic correlations were estimated employing LAVA *(Local Analysis of [co]Variant Association)*(^64^) across 7,351 LD blocks (∼1 Mb each, excluding the extended MHC region). For each trait pair, loci where both traits exhibited univariate genetic signal (FDR < 0.05) were subjected to bivariate testing, resulting in 20,582 tests; significance was defined by Bonferroni correction (*P*-values < 0.05/20,582 = 2.4×10^-6^).

### Multiple testing correction

For S-LDSC, we computed Benjamini-Hochberg (BH) FDR within each trait (342 tests per SUD, encompassing 57 cell types × 6 annotation categories) as the primary correction. FDR-significant results (FDR < 0.05) are reported as the main findings; nominally significant results (*P*-values < 0.05, FDR ≥ 0.05) are described as hypothesis-generating.

### Mendelian randomization

Bidirectional two-sample MR was performed between T2D (Suzuki et al., T2DGGI; GRCh38) and each of the four SUDs using the *TwoSampleMR* R package(^65^). Genome-wide instruments were selected at *P*-values < 5×10^-8^ and clumped for linkage disequilibrium (*r*^2^ < 0.001, 10 Mb window) using the European 1000 Genomes reference. Instrument strength was verified by computing per-SNP F-statistics; SNPs with F < 10 were excluded. For TUD, which reports per-SNP effective sample size as a Weight column rather than a fixed N, standard errors were derived as SE = 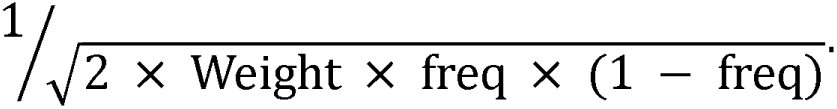 Five complementary MR methods were applied: inverse-variance weighted (IVW), MR-Egger, weighted median, weighted mode, and robust adjusted profile score (RAPS). Sensitivity analyses included the MR-Egger intercept test for directional pleiotropy and Steiger directionality filtering to confirm correct causal orientation. To account for multiple testing across 12 independent MR analyses (8 genome-wide + 4 cis), we applied BH-FDR using an adaptive primary estimator: IVW was the default; when the Egger intercept was significant (*P* < 0.05), indicating that IVW was biased by directional pleiotropy, the weighted median was used as the primary estimator for that analysis. FDR was computed across the 12 selected primary *P*-values.For drug-target (cis) MR at the *GLP1R* locus (chr6:38,916,557-39,179,018; GRCh38), instruments were selected within ±100 kb of the gene boundary at a relaxed threshold (*P*-values < 1 × 10^-5^, *r*^2^ < 0.1), following the Schmidt et al. framework(^66^).

### Pancreatic-SUD axis analyses

To evaluate the similarity between pancreatic and neural transcriptomic profiles, we performed sPLS-DA and DIABLO analysis using the *mixOmics* (^67^). To map directed cross-tissue signaling disruptions, we mapped SNPs via H-MAGMA(^68^) to genes that carried significant genetic burden (FDR ≤ 0.05), then integrated with the CellChat database to identify valid pancreatic ligand and neural receptor pairs within secreted signaling networks. We then quantified the aggregate genetic burden of these neuro-endocrine interactions via custom gene-set enrichment analyses in MAGMA, calculating pair-specific Z-scores to evaluate distinct pathway liabilities across SUDs.

### Developmental Trajectory and Module Burden analyses

We quantified aggregate risk by averaging gene Z-scores across pseudotime modules of cell-type-specific risk genes inferred by Slingshot(^69^) across hESC-derived hypothalamic lineages.

## RESULTS

### Distinct Genetic Architectures of Substance Use Disorders

To assess cell-type specific heritability, we applied S-LDSC(^49^) across all 59 human cell types (immune, metabolic, neuronal, and pluripotent lineages - **Table S1**), utilizing recent European population GWAS summary statistics for TUD, AUD, CanUD, and OUD (**Table S2**). Integrating ATAC-seq and 3D genomic maps (PCC, Hi-C) we defined six functional annotation categories per cell type (**Figure 1A**; detailed comparisons in **Supplementary Information and Figure S1-4**). We calculated partitioned heritability enrichment (**Figure S1**) and standardized effect size (*τ\**), applying per-trait BH-FDR correction (**Figure 1B**).

**Figure 1.**
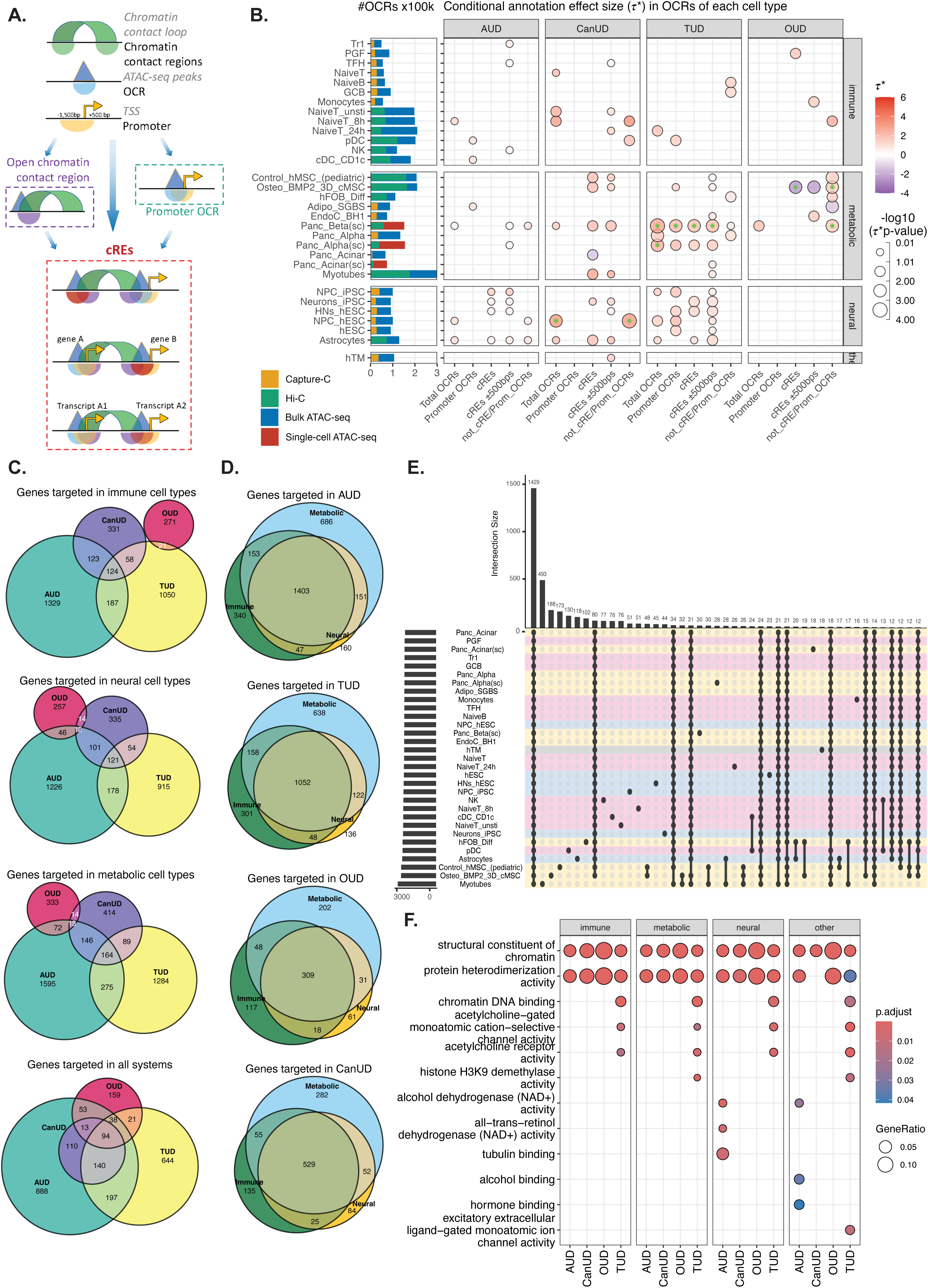
Systemic regulatory landscape and functional genomic architecture of SUDs. **A**. **Schematic of the functional annotation strategy** integrating ATAC-seq open chromatin regions (OCRs) with 3D genomic interaction maps (Hi-C, Promoter Capture-C) to define promoter-associated OCRs (Promoter OCRs) and distal putative cis-regulatory elements (cREs) that physically contact gene promoters. **B. Substance use disorders conditional effect size (***τ***\*) in annotation across diverse cell types:** Bar plots (left) depict total number of open chromatin regions (OCR x100,000) identified by bulk (blue) or single-cell (red) ATAC-seq and the proportion of those OCR contacting a gene promoter (putative cRE) as measured by Hi-C (green) or promoter capture-C (orange). The dot plots depict nominally significant enrichment (Enrichment *P*-values ≤0.05) within OCR annotations for each cell type across 4 SUDs as determined by S-LDSC analysis. The colors of the dots correspond to conditional effect sizes *τ*\* values, with dots featuring an asterisk indicating *τ*\* FDR ≤ 0.05. The size of the dots corresponds to *τ*\* *P*-values in-log10. **C. Cross-disorder gene overlap:** Venn diagrams showing the intersection of risk genes (mapped via H-MAGMA) shared among AUD, CanUD, TUD, and OUD within immune, neural, and metabolic tissue systems. **D. Cross-system gene overlap within disorders:** Venn diagrams illustrating the overlap of targeted genes across immune, metabolic, and neural systems for each specific SUD, highlighting the systemic distribution of genetic risk. **E. Cell-type specificity of risk genes:** UpSet plot visualizing the intersection size of mapped risk genes across individual cell types, displaying the number of unique and shared genes across the cellular panel. **F. Functional enrichment analysis:** Dot plot of top enriched Gene Ontology (GO) Molecular Function terms for the mapped risk genes, stratified by system and disorder. Dot size represents the gene ratio, and color indicates the Benjamini-Hochberg adjusted P-value.

After FDR correction, significant heritability enrichment was concentrated in TUD and CanUD (**Figure 1B, Table S3**). TUD showed FDR-significant enrichment (FDR < 0.05) in pancreatic β-cells (4.6-fold in Total OCRs, 4.9-fold in cREs), pancreatic α-cells (4.0-fold in Total OCRs), iPSC-derived neurons (2.4-fold in cREs ± 500 bp), and hypothalamic neurons (2.6-fold in cREs ± 500bp), spanning both metabolic and neural lineages across 8 cell-type-annotation combinations. CanUD enrichment was concentrated in hypothalamic neural progenitor cells (NPCs) (7.4-fold in not-cREs/Prom OCRs, FDR = 0.010; 5.8-fold in Total OCRs, FDR = 0.038). AUD and OUD did not survive FDR correction for enrichment, although nominal enrichment (*P*-values < 0.05) was observed across 14 and 8 cell types, respectively. For *τ\**, FDR-significant effects were observed for CanUD in hypothalamic NPCs (*τ\** = 3.45) and OUD in adipocytes (*τ\** = −4.06).

At the nominal level, broader patterns emerged. Total OCRs showed positive enrichment (>1) in 52 of 59 cell types for AUD, TUD, and CanUD, and 50 for OUD, though most did not survive FDR correction. Promoter OCRs exhibited high variability, with enrichment correlating with gene expression primarily in neural lineages. cREs and cREs ± 500 bp demonstrated the strongest enrichment, particularly in neural and pancreatic lineages, while not-cREs/Prom OCRs served as an accessible-but-dormant baseline.

### Neural Lineages

Across the three SUDs with nominal neural enrichment (AUD, CanUD, TUD), NPCs and excitatory neurons consistently exhibited the highest fold-enrichment scores (> 5-fold, *P*-values < 0.05), suggesting a developmental vulnerability that persists into neuronal maturity. NPC heritability was broadly distributed across genomic annotations, contrasting with the distal-specific cREs enrichment observed in neurons. CanUD showed an FDR-significant effect in hypothalamic NPCs (*τ* ≍* 3.5) nearly double that of any other SUD in this cellular setting. Conversely, TUD exhibited a pan-developmental profile, with FDR-significant enrichment in both iPSC neurons and hypothalamic neurons. AUD showed moderate *τ\** in NPCs, neurons and astrocytes. OUD risk showed no significant neural enrichment within the current cell-type panel. CanUD had the highest density of risk in astrocytes (*τ* ≍* 2.0), followed by TUD (*τ* ≍* 1.3) and AUD (*τ*\* *≍* 0.9).

### Metabolic cell types

A notable finding was the FDR-significant heritability enrichment for TUD in both pancreatic β-and α-cells across multiple annotation categories. CanUD exhibited a broad metabolic profile with nominally elevated *τ*\* values (*≍*2–3) across adipocytes, myotubes, osteoblasts, and *β*-cells, aligning with the endocannabinoid system’s systemic role in energy homeostasis. CanUD risk was specific to the endocrine pancreas, showing negative signals in exocrine acinar cells.

AUD showed a small positive *τ*\* in SGBS-adipocytes. OUD exhibited a unique bidirectional profile in metabolic and mesenchymal lineages: in *β*-cells, risk was enriched in non-looping “not-cREs/Prom OCRs”, while mesenchymal lineages displayed opposing signatures: osteoblasts showed positive *τ*\* in non-looping regions but negative enrichment in active cREs, whereas adipocytes exhibited negative *τ*\* in the non-looping regions.

### Immune cell types

CanUD and OUD showed the highest median immune *τ*\* (*≍* 2.0-2.2), significantly higher than AUD (*≈* 0.8), though these differences did not survive FDR correction. Within the B-cell lineage, OUD risk was nominally enriched in active cREs of PGF cells, while TUD risk was concentrated in non-looping OCRs of naive and germinal center B-cells. Additionally, CanUD and OUD displayed nominally specific enrichment within candidate poised enhancers (“not-cREs/Prom OCRs”) of stimulated T-cells, implying that vulnerability to these disorders lies in the immune response to stress rather than resting function.

Monocyte enrichment (cREs±500bp) indicated that OUD variants disrupt chromatin context rather than core motifs. This unique signature, absent in the other three SUDs, implicates monocytes as a specific OUD cell type.

While the overall immune signal was lower, AUD revealed specific signatures: Promoter OCRs in Dendritic cells (pDC, cDC) and cREs in NK/T-regs (TR1, TFH).

### Trabecular meshwork (hTM) cells

Human trabecular meshwork cells exhibited a nominally significant heritability enrichment for CanUD within cREs ±500bp (*τ*\*=1.44).

### Implicating SUD effector genes

To assign these non-coding signals to their corresponding effector genes, we employed H-MAGMA(^68^) utilizing our cell-specific 3D chromatin contacts and OCRs to capture distal enhancer interactions. We implicated 5,651 candidate risk genes (FDR < 0.05) across our cellular library (**Table S4**), 4,557 candidate genes in metabolic cell types, 3,694 candidate genes in immune cell types, and 3,349 candidate genes in neural lineages (**Figure 1C, Figure S5**).

Across all tissue types, AUD and TUD displayed the broadest overlap, sharing 275 candidate genes in metabolic tissues. Extensive cross-tissue pleiotropy (**Figure 1D**) revealed that the largest proportion of candidate risk genes was shared across immune, metabolic, and neural lineages (1,403 for AUD and 1,052 for TUD). While OUD showed a more restricted but metabolic-concentrated pattern, the largest intersection set represented ubiquitous genes active across all 31 cell types (**Figure 1E**).

Gene Ontology analysis revealed universal enrichment for the categories of ‘structural constituent of chromatin’ and ‘protein heterodimerization activity’ across all four SUDs (**Figure 1F**). TUD and CanUD converged on rapid neurotransmission (acetylcholine receptors, ion channels). AUD was characterized by metabolic enzymatic processes, ‘alcohol dehydrogenase (NAD+) activity’ and ‘retinol binding’.

However, distinct signatures of cell-type specificity remained. Notably, pancreatic lineages frequently clustered with broad systemic sets, suggesting that the pancreatic signal in SUDs is driven by a broadly active regulatory machinery that is functionally relevant in both neurons and secretory metabolic cells.

### Shared genetic architecture between SUDs and type 2 diabetes

We investigated the observed pancreatic-SUD axis using estimated genome-wide genetic correlations (r_g_) between SUDs and a range of pancreatic and glycemic traits (**Figure 2A**). TUD and CanUD showed significant positive correlations with T2D and HbA1c concentration. Conversely, AUD was correlated positively with hypoglycemia but negatively with T2D. There were no correlations with type 1 diabetes.

**Figure 2.**
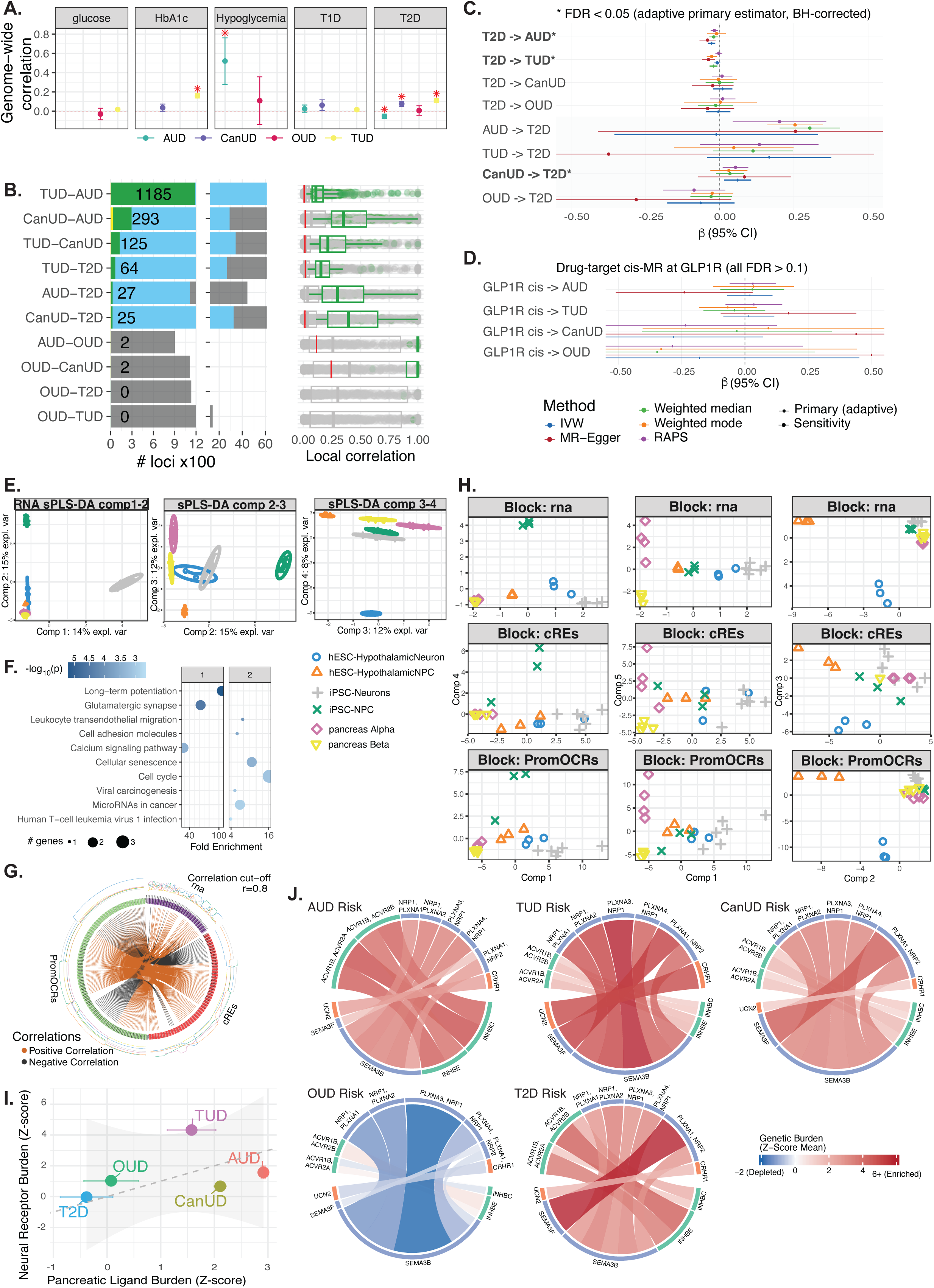
Multi-omic Integrations of SUDs and Metabolic Traits. **A. Genome-wide correlation** between SUDs and metabolic indicators. **B. Local genetic correlation:** Barplot (left) shows numbers of loci tested (grey), found correlated (blue), and reached a significant FDR≤0.05 (green) for each pair-wise of disorders; yellow bars are the number of loci where the CI-95 of correlation between the pair reached 1. Scatter plot (right) showing distributions of correlations across loci for each pair of disorders, green boxplots were significant correlated loci, red lines indicate the correlation level at genome-wide scale. **C-D. Forest plot showing causal effect estimates (***β***, 95% CI) from bidirectional two-sample Mendelian randomization between T2D and SUDs: C. Genome-wide analyses** (8 directions), 5 MR methods per analysis: inverse-variance weighted (IVW, blue), MR-Egger (red), weighted median (green), weighted mode (orange), and robust adjusted profile score (RAPS, purple), diamond markers (⍰) denote the pre-specified primary estimator for each analysis, circle markers (●) denote sensitivity methods; Benjamini-Hochberg FDR was computed across the 12 primary P-values; bold labels with asterisks (*) indicate FDR < 0.05: T2D→AUD (IVW *β*=-0.027, FDR = 1.9 × 10-4), T2D→TUD (weighted median *β* = - 0.020, FDR = 0.013), and CanUD→T2D (IVW *β* = 0.061, FDR = 0.036). For AUD→T2D, IVW was null (*P* = 0.95) but the weighted median (P = 4.8 × 10^-9^) and weighted mode (P = 2.6 × 10^-6^) were highly significant, suggesting outlier-driven IVW bias (Egger intercept P = 0.36, no directional pleiotropy detected). **D. Drug-target cis-MR** using T2D-associated variants within ±100 kb of GLP1R (chr6:38,916,557-39,179,018; GRCh38) as instruments (5-15 SNPs per analysis; P < 1 × 10-5, r² < 0.1, the dashed vertical line indicates *β* = 0 (no causal effect). **E. Cell types projection from sPLS-DA analysis** performed using total expression profiles of 6 cell types into 4 optimized components space; ellipse represents 95% confidence intervals around and colored by each sample class; 1^st^ component separated iPSC-derived neurons, 2^nd^ component separated iPSC-derived NPC, 3^rd^ component separated 2 pancreatic cell types and 4^th^ component separated hESC-derived hypothalamus neurons from the rest but all combination failed to separate pancreatic cells from neural cell types. **F. Pathways enrichment** from a few genes drove the separation of components 2 and 4 from panel A. **G. Multi-omic correlation circle** plot represents the correlations greater than 0.8 between variables of different types, represented on the side quadrants, internal connecting lines show the positive (negative) correlations, the outer lines show the expression levels of each variable in each cell type. **H. Multiblock DIABLO analysis** sample plot performed using 3 blocks of RNA-seq profiles and ATAC-seq profiles from Promoter OCRs and cREs within 114 loci displayed significant correlation between T2D and SUDs; the representative 3 combinations of 5 optimized components showed the most separation power, colored similar to panel A: 1^st^ component effectively separated primary lineages (neural progenitors, differentiated neurons, and pancreatic cells), 2^nd^ component focused on distinguishing hESC-derived neural progenitor cells, 3^rd^ component characterized hESC-derived hypothalamic neurons, 4^th^ component separated iPSC-derived neural progenitor cells, 5^th^ component differentiated pancreatic *α*-and *β*-cells and neural cells; similar to panel E, still failed to distinguish pancreatic cell from neural cell types. **I**. **Genetic burden (Z-score Mean)** of ligands in the pancreas versus receptors in neural tissue. Points are colored by trait (T2D, OUD, TUD, AUD, CanUD). **J. Neural-pancreatic signaling chords plots** illustrating specific ligand-receptor interactions across different risk profiles, color intensity represents the Genetic Burden (Z-score Mean), showing enrichment or depletion of specific signaling pathways in each disorder.

Because global correlations average out opposing signals, we used LAVA (^64^) to investigate genetic regions associated with each SUD and T2D, which revealed 1,550 loci with Bonferroni-significant bivariate local correlations (*P*-values < 0.05/20,582) across the 10 pairwise SUD-T2D combinations (**Figure 2B**, **Table S5**), of which 114 were shared between T2D and at least one SUD. The top locus, chr4:99187511-99411908, connected T2D to AUD (r_g_=0.8). TUD shared 64 loci with T2D (r_g_=0.04-0.6), while AUD shared 27 (r_g_=0.05-0.8) and CanUD shared 25 (r_g_=0.1-0.8) loci.

### Mendelian randomization validates a causal T2D-AUD relationship

To provide orthogonal validation of the pancreatic-SUD axis, we performed bidirectional two-sample MR between T2D and each SUD (**Figure 2C, Table S6**). Three of 12 analyses survived BH-FDR correction (FDR < 0.05) using the pre-specified adaptive primary estimator (see Methods).

In the forward direction, genetic liability for T2D was protective against AUD (IVW: *β* =-0.027, SE = 0.006, *P* = 1.6 × 10^-5^; 463 instruments), FDR = 1.9 × 10^-4^, consistent across four of five methods: MR-Egger (*β* =-0.040, *P* = 0.004), weighted median (*β* = - 0.020, *P* = 0.008), RAPS (*β* =-0.018, *P* = 0.006), and weighted mode (*β* =-0.010, *P* = 0.51). The MR-Egger intercept was not significant (*P* = 0.28), indicating no directional pleiotropy, and Steiger filtering confirmed correct causal direction (*P* = 2.9 × 10^-214^).

Toward TUD, T2D genetic liability did not achieve significance (IVW *P* = 0.13; 476 instruments), FDR = 0.013, but the MR-Egger intercept was significant (*P* = 0.003), indicating directional pleiotropy contaminating the IVW estimate. The adaptive rule selected the weighted median as the primary estimator (*β* =-0.020, *P* = 0.002), which was corroborated by weighted mode (*β* =-0.026, *P* = 0.003), and MR-Egger (*β* =-0.037, *P* = 0.0009), converged on a significant protective effect of T2D liability on TUD. T2D showed no significant causal effect on CanUD (IVW *P* = 0.54) or OUD (IVW *P* = 0.82).

In the reverse direction CanUD showed a significant risk-increasing effect on T2D (IVW: *β* = 0.061, *P* = 0.009; RAPS *P* = 0.015; 17 instruments), FDR = 0.036 and no evidence of pleiotropy. TUD, AUD, and OUD did not show FDR-significant reverse causal effects on T2D by IVW, though AUD liability toward T2D exhibited a notable discordance: while IVW was null (*P* = 0.95; 49 instruments) and the Egger intercept did not detect pleiotropy (*P* = 0.36), the weighted median (*P* = 4.8 × 10^-9^) and weighted mode (*P* = 2.6 × 10^-6^) strongly suggested a positive causal effect, possibly reflecting outlier instrument bias in the IVW estimate.

Drug-target (cis) MR at the *GLP1R* locus (**Figure 2D**) did not reach significance for any SUD (AUD: *P* = 0.67; TUD: *P* = 0.77; CanUD: *P* = 0.12; OUD: *P* = 0.58, all FDR > 0.1), suggesting that the causal metabolic-SUD relationship operates through pathways broader than GLP-1 receptor signaling alone.

### Functional convergence of neural and pancreatic regulation at shared loci

We next investigated whether T2D-SUD risk overlap is mediated by shared conserved regulatory mechanisms. Principal component analysis (PCA) confirmed that neural and pancreatic lineages remained globally distinct (**Figure S6**). However, accessing the top 10% specifically expressed genes revealed 4,717 candidate genes shared across various combinations of these cell types (**Figure S7**). Although the universal intersection was small (294 genes), this overlap suggests that functions across these otherwise distinct lineages are conserved.

Sparse Partial Least Squares discriminant analysis (sPLS-DA) from mixOmics (^67^) (**Figure 2E-F, Figure S8**) resolved cellular identity. While components 1 and 2 separated lineages, Component 3 clustered *β*-cells closely with hESC-hypothalamic neurons and iPSC-NPCs. This convergence was enriched for primary ciliary genes like *DCAF6*, providing evidence for a pancreatic-SUD axis of shared regulatory architecture.

Focusing on the 114 shared loci, we used the ‘Data Integration Analysis for Biomarker discovery using Latent cOmponents’ (DIABLO) framework, also from mixOmics(^67^), to integrate RNA-seq and ATAC-seq (promoter OCRs and cREs). We observed high correlation (>0.8) between gene expression and chromatin accessibility within these risk regions (**Figure 2G-H, Figure S9**). This observation suggests that shared genetic risk operates through a highly coordinated regulatory logic common to both cell types, which the model parsed into distinct biological processes (**Figure S10**). Mapping regulatory elements revealed distinct mechanisms (**Figure S11**). AUD-T2D converged on metabolic enzymes (alcohol/retinol dehydrogenase), TUD-T2D on cytoskeletal dynamics and ion transport (microtubules, potassium channels), and CanUD-T2D on lipid signaling (diacylglycerol kinase) (**Figure S12**).

While globally distinct, neural and pancreatic lineages converge at T2D-SUD loci via shared regulatory architectures for secretion, metabolism, and membrane dynamics.

### Differential disruption of shared neuro-endocrine signaling axes

Given our observations of shared intracellular machinery, we tested for potential direct signaling networks between neurons and *β*-cells. Hypothesizing a functional link, we built a ‘sender-receiver’ model using the *CellChat* (v2.2.0) ligand-receptor repository to map our observed tissue-specific risk genes (**Signaling Inference and Network Construction – Supplementary Method**) We filtered pancreatic risk genes for ligands and neural genes for receptors, defining putative impacted interactions, where both carry significant genetic burden. This identified candidate neuro-endocrine axes with predicted genetic vulnerabilities in both signal generation and reception.

Examination of the aggregate genetic burden across the pancreatic-SUD axis (**Figure 2I**), T2D clustered near the origin (*Z≈*0), establishing a baseline. Against this, TUD risk was concentrated in neuronal receptors (*Z≈*4.5), suggesting a potential a risk defect in neural sensing. Conversely, AUD risk skewed toward pancreatic ligands (*Z≈*3), consistent with predicted defective signal output. OUD showed minimal burden, aligning with its primary risk in upstream chromatin regulation.

CellChat analyses revealed that divergence correlates with differential genetic burden at opposite termini of shared neuro-endocrine axes (**Figure 2J**). TUD exhibited the strongest risk enrichment within the structural (SEMA3*→*NRP1/PLXNA1) and stress (UCN2*→*CRHR1) axes. In contrast, AUD and T2D shared the metabolic activin axis (INHBC/INHBE*→*ACVR1B). TUD and CanUD specifically overlapped with T2D via the semaphorin guidance pathway (SEMA3F*→* PLXNA1). Consequently, T2D displayed a dual signature of shared liability: overlapping with AUD via activin-mediated signaling and with TUD/CanUD via the predicted semaphorin guidance pathway. OUD lacked significant enrichment in these axes.

These inferred signaling disruptions represent candidate mechanisms that warrant *in vivo* validation in the future.

### Comparison with psychiatric comorbidities

To investigate whether the observed SUD-pancreatic enrichments are merely an epiphenomenon that driven by the known 3D architectural similarities between neurons and pancreatic endocrine cells, we evaluated the cell-type-specific enrichment profiles of other highly heritable psychiatric disorders. Genome-wide correlation (**Figure 3A**) confirmed pervasive and significant positive associations between SUDs and major psychiatric conditions, most notably attention-deficit/hyperactivity disorder (ADHD), schizophrenia (SCZ), major depressive disorder (MDD), and post-traumatic stress disorder (PTSD). Subsequent S-LDSC analysis confirmed that ADHD, SCZ, and bipolar disorder (BIP) also showed significant enrichment (*P*-values < 0.05) across immune, metabolic, and neural lineages. (**Figure 3B**), this is not a universal rule. Eating Disorders (ED) demonstrated significant genetic enrichment across neural lineages with a complete absence of signal in pancreatic cells. Conversely, traits such as Anxiety (ANX) and Parkinson’s Disease (PD) exhibited the exact opposite pattern: significant enrichment within pancreatic cells (Panc_Alpha(sc) and Panc_Beta(sc), respectively) but a notable absence of enrichment across all assessed neural lineages. This dissociation confirms that metabolic and neural enrichments vary independently across disorders.

**Figure 3.**
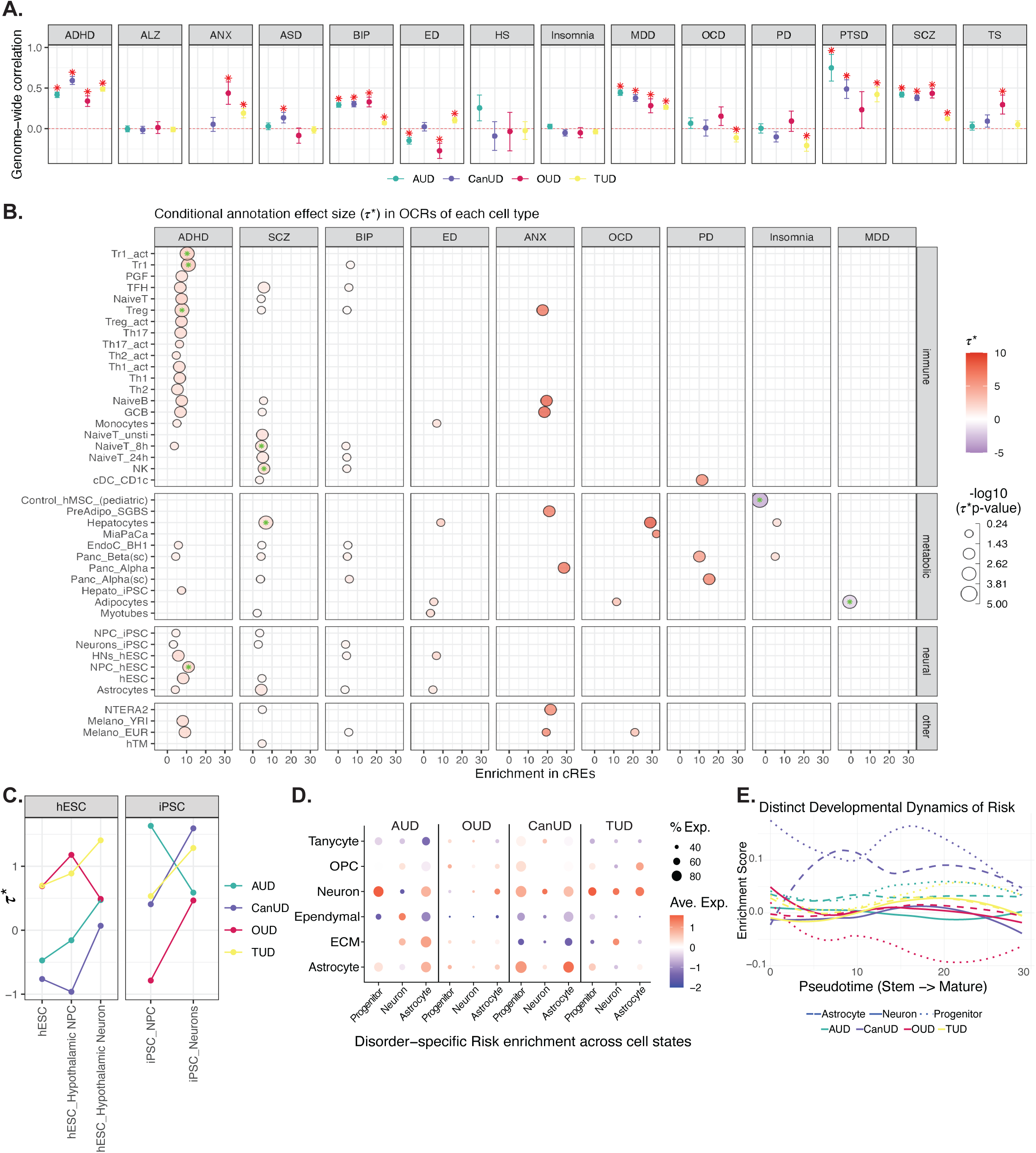
Genetic overlap and cellular enrichment of SUDs across neuropsychiatric traits. **A. Genome-wide correlations** between 4 SUDs and panels of 14 major psychiatric conditions, with red asterisks above dots indicate significant correlation *P*-values ≤0.05. **B. Cell-Type Specific Enrichment in cREs:** Dot plots depicting conditional effect sizes *τ*\* for SUDs across each cell type but focus only on cREs, x-axis is Enrichment scale. **C. Lineage-specific enrichment** line graphs showing the change in *τ*\* across developmental lineages for both human embryonic stem cells (hESC) and induced pluripotent stem cells (iPSC), tracks the genetic risk enrichment as cells differentiate from progenitors into mature neurons or hypothalamic neurons. **D. Disorder-specific risk enrichment** bubble plot characterizing SUDs across different brain cell states, includes Progenitors, Neurons, Astrocytes, Ependymal cells, and more; size of the bubble indicates the percentage of expression, while the color represents the average expression level. **E.** Line chart mapping the **Enrichment Score against Pseudotime** show the genetic risk for SUDs peaks at different stages of cellular maturation.

### Neural lineage specificity and development

While OUD was characterized by a widespread metabolic/immune signature, other SUDs converged on a distinct neural signature. Isolating the neural compartment suggests that their shared liability is not driven by signaling function of differentiated neurons but rooted in the chromatin landscape of early neurogenesis.

To disentangle developmental wiring from mature neuronal function, we stratified neural heritability across differentiation trajectories (Stem Cell*→*Progenitor*→*Neuron) (**Figure 3C**). We then validated cellular specificity by constructing functional modules with risk genes identified through H-MAGMA for each cell type and SUD (**Supplementary Methods**), assessing their relative gene expression patterns within immature hypothalamic single-cell data (ages 4-14 years)(^70^) (**Figure 3D**). Mapping was restricted to hESC-derived hypothalamic lineages to ensure regional consistency. Consistent with primary S-LDSC findings, OUD risk modules were almost absent in the childhood dataset. While transient signals *τ*\* appeared in progenitors, they did not persist.

TUD displayed a neuron-centric risk architecture. Regardless of whether candidate risk genes were identified in progenitor, neuron, or astrocyte modules during differentiation, their expression mapped preferentially to the immature neuron cluster. AUD exhibited a split profile with risk genes from the *AUD_Progenitor* module highly expressed in immature neurons, while risk genes from the *AUD_Astrocyte* module expressed specifically in astrocytes. CanUD displayed a broad signature where all risk modules were expressed across both neurons and astrocytes.

To reconstruct the temporal emergence of genetic liability, we modeled risk scores along a pseudotime differentiation trajectory rooted in hypothalamic stem cells, revealing two distinct mechanisms (**Figure 3E**). *TUD_Neuron* modules all remained low or negative in early stages before rising toward terminal differentiation. In contrast, AUD risk follows a constitutive trajectory, maintaining high enrichment levels from the earliest stem-like states through to differentiation.

## DISCUSSION

We sought to identify the cellular contexts that mediate SUD etiology by examining epigenetic enrichment patterns across diverse cell types. Building on the growing recognition of non-neuronal mediators in addiction (^71–77^), we integrated GWAS summary statistics with cell-type-specific 3D-genomic maps(^70, 78–84^) to map this multi-systemic architecture. Our results expand the traditional neuro-centric model by revealing that genetic risk for SUDs is distributed across a pancreatic-SUD axis and distinct immune lineages. While we confirmed the expected role of cortical neurons and neural progenitor cells (NPCs) the heritability enrichment in pancreatic *α*-and *β*-cells across three disorders suggests that the physiological drive for substance use is also partially encoded in the same regulatory machinery governing energy homeostasis.

Importantly, the pancreatic-SUD axis receives orthogonal validation from Mendelian randomization. Genetic liability for T2D is causally protective against AUD, observed robustly across multiple MR methods and free of directional pleiotropy. For TUD, although IVW was not significant due to directional pleiotropy, the pleiotropy-robust estimators (weighted median, weighted mode and MR-Egger) converged on a significant protective effect in the same direction. In the reverse direction, CanUD liability increased T2D risk. These protective directions are consistent with the negative AUD-T2D and positive TUD/CanUD-T2D genetic correlations we observed, and align with epidemiological evidence that metabolic interventions can reduce alcohol and tobacco consumption (46, 47). The MR findings establish that the SUD-T2D genetic overlap reflects shared causal architecture rather than confounding or pleiotropic artifact.

This pancreatic-SUD axis reflects an evolutionary conservation where insulin-producing neurons phylogenetically precede pancreas, *β*-cells retain a neuronal-like gene expression program, and chromatin landscape is enabled by the depression of polycomb proteins during organogenesis(^85–88^). Our data indicate that SUDs potentially co-opt these shared functional modules, but through distinct mechanisms. AUD exhibits a metabolic sender profile driven by putative pancreatic ligand defects (activin), whereas TUD displays a neural receiver profile characterized by predicted receptor dysfunction (SEMA3, UCN). By demonstrating that neural and pancreatic enrichments vary independently across other disorders (e.g., ED versus ANX/PD), we confirm that the metabolic signals observed in SUDs are distinct etiological features rather than artifacts of shared 3D architecture. Importantly, because these ligand-receptor models are inferred from the *in silico* integration of genetic risk profiles with curated CellChat databases, they represent candidate signaling networks that will require future *in vivo* validation to confirm active systemic neuroendocrine crosstalk.

Chronic substance exposure precipitates glucose dysregulation and diabetes risk(^89–91^). Distinct mechanisms drive this pathology: alcohol toxicity induces *β*-cell dysfunction and glucagon dysregulation(^92–94^); opioids directly modulate *α*-and *β*-cell secretion to disturb glucose homeostasis(^95, 96^); and cannabinoids activate CB1 receptors on these cells to impair insulin while stimulating glucagon(^97–99^). Furthermore, nicotine use links to the pancreas via the T2D-associated transcription factor TCF7L2, which has been shown to regulate intake through habenular nicotinic receptors and influences blood glucose via the autonomic nervous system(^100^).

Beyond the pancreas, our extended analysis of psychiatric traits suggested a divergence in the genetic architecture of addiction, differentiating early neurodevelopmental enrichment profiles from metabolic and immune ones. TUD and CanUD align more closely with the patterns of ADHD and schizophrenia, evidenced by robust signals in fetal NPCs and hypothalamic neurons. While TUD mirrors the mature neuronal enrichment seen in ADHD and SCZ, CanUD risk is highly concentrated in the developing hypothalamus. This temporal ordering raises the possibility that metabolic and immune dysfunctions in these specific SUDs may manifest secondary to earlier neural vulnerabilities.

Consistent with well-documented SUD immunomodulation, our observations align with cytokine dysregulation, CNS inflammation, and impaired immune function characteristic of chronic alcohol consumption(^101–105^). OUD was uniquely characterized by heritability enrichment in the innate immune system (monocytes/macrophages), aligning with the capacity of opioids to bind toll-like receptor 4 (TLR4) and drive sterile inflammation. This dissociates OUD from the adaptive immune (T-cell) involvement seen in AUD, CanUD, and schizophrenia. Collectively, these findings suggest that OUD is a disorder of cellular adaptation, driven by a failure of peripheral mesenchymal and innate immune tissues to manage systemic stress, rather than an intrinsic defect in neurotransmission.

Recognizing these distinct architectures opens avenues for precision medicine. The convergence of SUDs and metabolic disorders suggests that repurposing metabolic therapeutics could be effective. Glucagon-like peptide-1 (GLP-1) receptor agonists (e.g., semaglutide) offer a potent trans-diagnostic intervention by structurally bridging the sender-receiver gap identified in the pancreatic-SUD axis. By targeting GLP-1 receptors expressed(^106–108^) in key reward-processing centers like the ventral tegmental area and nucleus accumbens(^109–111^), these agents modulate dopaminergic signaling to attenuate drug-seeking behavior while simultaneously restoring the metabolic homeostasis disrupted in addiction(^48^). This dual mechanism directly addresses the systemic pathology of SUDs: it stabilizes the ‘sender’ (pancreatic metabolic output) and rectifies the ‘receiver’ (neural reward sensitivity), effectively treating the metabolic dysregulation that reciprocally drives addiction liability. Our drug-target cis-MR at the *GLP1R* locus was null for all four SUDs, suggesting that the therapeutic potential of GLP-1 agonists for addiction may operate through downstream metabolic pathways rather than direct GLP1R-mediated effects, or that the cis-MR analysis was underpowered for these outcomes. Notably, our immune results argue for a more nuanced interpretation. While GLP-1 agonists may address the metabolic-neural interface, immune-based therapies must be tailored: targeting the innate inflammatory response (anti-TLR4) may be effective for OUD, while modulating immune tolerance (Treg function) may be more relevant for the adaptive immune dysregulation observed in AUD and CanUD. Thus, although increasingly, GLP-1 receptor agonists are viewed as potential therapies for a wide variety of medical and psychiatric disorders, it appears likely that the magnitude of their efficacy, and importantly their adverse event profiles, will depend upon and be predicted by a complex array of etiological factors that differentiate the target disorders.

Several limitations should be considered. Our analyses used European-ancestry GWAS exclusively; generalizability to other populations requires future replication in cross-ancestry cohorts. While our use of cell-type-specific chromatin maps provides higher resolution than broad tissue annotations, integrate across diverse library protocols and cell culture conditions, though down-sampling sensitivity analysis demonstrated that enrichment patterns remained consistent at uniform sequencing depths (**Figure S4**). AUD and OUD did not survive FDR correction for heritability enrichment; we report nominal patterns for these disorders as hypothesis-generating, noting that pancreatic involvement for AUD is independently supported by MR evidence and the well-characterized effects of alcohol on *β*-cell function. We acknowledge that local genetic correlations between SUDs and metabolic traits are often modest. However, the biological insight derives from the aggregation of these signals into coherent pathways. Our LAVA and sPLS-DA analyses demonstrated that these loci are not random but converge on specific regulatory elements active in both lineages. Future work is needed to map the’missing’ heritability of OUD onto peripheral axes, specifically the adipocyte and osteoblast reservoirs identified here, to pinpoint the somatic cell states driving opioid liability.

## CONCLUSION

Cumulative evidence(^112–114^) suggests that SUDs are not merely brain diseases. From a genetic perspective, our findings support the concept that the risk architecture for addiction is multi-systemic: it is wired into the developing hypothalamus in CanUD and TUD, pancreatic endocrine cells for TUD, and uncoupled from metabolic signaling in AUD. Mendelian randomization provides causal evidence linking T2D genetic liability to reduced AUD risk, offering a mechanistic foundation for the emerging clinical utility of GLP-1 receptor agonists in addiction medicine. Moving forward, therapeutic strategies should address the distinct metabolic, immune, and neural vulnerabilities that define each disorder.

## Data Availability

All data produced in the present study are available upon reasonable request to the authors

## ACKNOWLEDGEMENTS

S.F.A.G. is the Daniel B. Burke Endowed Chair for Diabetes Research. H.R.K is the Karl E. Rickels Professor of Psychiatry.

## CRediT author statement

**Khanh B. Trang**: Conceptualization, Methodology, Formal analysis, Investigation, Data Curation, Writing - Original Draft, Visualization. **Alessandra Chesi**: Formal analysis, Investigation, Data Curation, Writing-Reviewing and Editing. **Sylvanus Toikumo**: Resources, Data Curation, Writing-Reviewing and Editing. **James A. Pippin**: Resources. **Matthew C. Pahl**: Methodology, Writing-Reviewing and Editing. **Joan M. O’Brien**, **Laufey T. Amundadottir**, **Kevin M. Brown**, **Wenli Yang**, **Jaclyn Welles**, **Dominic Santoleri**, **Paul M. Titchenell**, **Patrick Seale**, **Babette S. Zemel**, **Yadav Wagley**, **Kurt D. Hankenson**, **Klaus H. Kaestner**, **Stewart A. Anderson**: Resources. **Matthew S. Kayser**: Funding acquisition; Writing, Reviewing and Editing. **Andrew D. Wells**: Resources; Writing, Reviewing and Editing. **Henry R. Kranzler**: Supervision; Funding acquisition; Writing, Reviewing and Editing. **Rachel L. Kember**: Supervision; Funding acquisition; Writing, Reviewing and Editing. **Struan F.A. Grant**: Conceptualization; Supervision; Funding acquisition; Writing, Reviewing and Editing

## TABLE LEGENDS

Table S1. Data resources of 59 cell types

Table S2. 4 substance use disorders and other psychiatric traits GWAS information

Table S3. S-LDSC results

Table S4. Implicated genes from H-MAGMA Table S5. LAVA results

Table S6. MR results and diagnostics

## REFERENCES

1. WHO (2018): Global status report on alcohol and health 2018.

2. SAMHSA (2022): National Survey on Drug Use and Health.Table 2.25—Alcohol use in lifetime: among people aged 12 or older; by age group and demographic characteristics, 2021 and 2022.

3. Esser MB SA, Liu Y, Naimi TS Deaths from Excessive Alcohol Use — United States, 2016–2021. MMWR and Morbidity and Mortality Weekly Report, pp 154–161.

4. WHO (2021): WHO global report on trends in prevalence of tobacco use 2000-2025.

5. Ahmad FB CJ, Rossen LM, Sutton P (2021): Provisional drug overdose death counts. National Center for Health Statistics: National Vital Statistics System.

6. Hasin DS, Saha TD, Kerridge BT, Goldstein RB, Chou SP, Zhang H, et al. (2015): Prevalence of Marijuana Use Disorders in the United States Between 2001-2002 and 2012-2013. JAMA Psychiatry. 72:1235–1242.

7. WHO (2016): The health and social effects of nonmedical cannabis use.

8. Lopez-Quintero C, Pérez de los Cobos J, Hasin DS, Okuda M, Wang S, Grant BF, et al. (2011): Probability and predictors of transition from first use to dependence on nicotine, alcohol, cannabis, and cocaine: results of the National Epidemiologic Survey on Alcohol and Related Conditions (NESARC). Drug Alcohol Depend. 115:120–130.

9. Winters KC, Lee CY (2008): Likelihood of developing an alcohol and cannabis use disorder during youth: association with recent use and age. Drug Alcohol Depend. 92:239–247.

10. Prom-Wormley EC, Ebejer J, Dick DM, Bowers MS (2017): The genetic epidemiology of substance use disorder: A review. Drug Alcohol Depend. 180:241–259.

11. Heath AC, Bucholz KK, Madden PA, Dinwiddie SH, Slutske WS, Bierut LJ, et al. (1997): Genetic and environmental contributions to alcohol dependence risk in a national twin sample: consistency of findings in women and men. Psychol Med. 27:1381–1396.

12. Kendler KS (2001): Twin studies of psychiatric illness: an update. Arch Gen Psychiatry. 58:1005–1014.

13. Sullivan PF, Kendler KS (1999): The genetic epidemiology of smoking. Nicotine Tob Res. 1 Suppl 2:S51–57; discussion S69-70.

14. Agrawal A, Verweij KJ, Gillespie NA, Heath AC, Lessov-Schlaggar CN, Martin NG, et al. (2012): The genetics of addiction-a translational perspective. Transl Psychiatry. 2:e140.

15. Agrawal A, Lynskey MT (2006): The genetic epidemiology of cannabis use, abuse and dependence. Addiction. 101:801–812.

16. Verweij KJ, Zietsch BP, Lynskey MT, Medland SE, Neale MC, Martin NG, et al. (2010): Genetic and environmental influences on cannabis use initiation and problematic use: a meta-analysis of twin studies. Addiction. 105:417–430.

17. Deak JD, Johnson EC (2021): Genetics of substance use disorders: a review. Psychological Medicine. 51:2189–2200.

18. Tsuang MT, Lyons MJ, Meyer JM, Doyle T, Eisen SA, Goldberg J, et al. (1998): Co-occurrence of abuse of different drugs in men: the role of drug-specific and shared vulnerabilities. Arch Gen Psychiatry. 55:967–972.

19. Kendler KS, Myers J, Prescott CA (2007): Specificity of genetic and environmental risk factors for symptoms of cannabis, cocaine, alcohol, caffeine, and nicotine dependence. Arch Gen Psychiatry. 64:1313–1320.

20. Kendler KS, Prescott CA, Myers J, Neale MC (2003): The structure of genetic and environmental risk factors for common psychiatric and substance use disorders in men and women. Arch Gen Psychiatry. 60:929–937.

21. Johnson EC, Chang Y, Agrawal A (2020): An Update on the Role of Common Genetic Variation Underlying Substance Use Disorders. Current Genetic Medicine Reports. 8:35–46.

22. Hancock DB, Markunas CA, Bierut LJ, Johnson EO (2018): Human Genetics of Addiction: New Insights and Future Directions. Current Psychiatry Reports. 20:8.

23. Edenberg HJ, McClintick JN (2018): Alcohol Dehydrogenases, Aldehyde Dehydrogenases, and Alcohol Use Disorders: A Critical Review. Alcohol Clin Exp Res. 42:2281–2297.

24. Gelernter J, Kranzler HR, Sherva R, Almasy L, Koesterer R, Smith AH, et al. (2014): Genome-wide association study of alcohol dependence:significant findings in African-and European-Americans including novel risk loci. Molecular Psychiatry. 19:41–49.

25. Kranzler HR, Zhou H, Kember RL, Vickers Smith R, Justice AC, Damrauer S, et al. (2019): Genome-wide association study of alcohol consumption and use disorder in 274,424 individuals from multiple populations. Nature Communications. 10:1499.

26. Walters RK, Polimanti R, Johnson EC, McClintick JN, Adams MJ, Adkins AE, et al. (2018): Transancestral GWAS of alcohol dependence reveals common genetic underpinnings with psychiatric disorders. Nat Neurosci. 21:1656–1669.

27. Clarke TK, Adams MJ, Davies G, Howard DM, Hall LS, Padmanabhan S, et al. (2017): Genome-wide association study of alcohol consumption and genetic overlap with other health-related traits in UK Biobank (N=112 117). Molecular Psychiatry. 22:1376–1384.

28. Gelernter J, Sun N, Polimanti R, Pietrzak RH, Levey DF, Lu Q, et al. (2019): Genome-wide Association Study of Maximum Habitual Alcohol Intake in >140,000 U.S. European and African American Veterans Yields Novel Risk Loci. Biological Psychiatry. 86:365–376.

29. Liu M, Jiang Y, Wedow R, Li Y, Brazel DM, Chen F, et al. (2019): Association studies of up to 1.2 million individuals yield new insights into the genetic etiology of tobacco and alcohol use. Nature Genetics. 51:237–244.

30. Sanchez-Roige S, Fontanillas P, Elson SL, Gray JC, de Wit H, Davis LK, et al. (2019): Genome-wide association study of alcohol use disorder identification test (AUDIT) scores in 20 328 research participants of European ancestry. Addict Biol. 24:121–131.

31. Xu K, Kranzler HR, Sherva R, Sartor CE, Almasy L, Koesterer R, et al. (2015): Genomewide Association Study for Maximum Number of Alcoholic Drinks in European Americans and African Americans. Alcohol Clin Exp Res. 39:1137–1147.

32. Zhou H, Rentsch CT, Cheng Z, Kember RL, Nunez YZ, Sherva RM, et al. (2020): Association of OPRM1 Functional Coding Variant With Opioid Use Disorder: A Genome-Wide Association Study. JAMA Psychiatry. 77:1072–1080.

33. Sanchez-Roige S, Palmer AA, Fontanillas P, Elson SL, Adams MJ, Howard DM, et al. (2019): Genome-Wide Association Study Meta-Analysis of the Alcohol Use Disorders Identification Test (AUDIT) in Two Population-Based Cohorts. Am J Psychiatry. 176:107–118.

34. Toikumo S, Jennings MV, Pham BK, Lee H, Mallard TT, Bianchi SB, et al. (2024): Multi-ancestry meta-analysis of tobacco use disorder identifies 461 potential risk genes and reveals associations with multiple health outcomes. Nat Hum Behav.

35. Saunders GRB, Wang X, Chen F, Jang S-K, Liu M, Wang C, et al. (2022): Genetic diversity fuels gene discovery for tobacco and alcohol use. Nature. 612:720–724.

36. Levey DF, Galimberti M, Deak JD, Wendt FR, Bhattacharya A, Koller D, et al. (2023): Multi-ancestry genome-wide association study of cannabis use disorder yields insight into disease biology and public health implications. Nat Genet. 55:2094–2103.

37. Demontis D, Rajagopal VM, Thorgeirsson TE, Als TD, Grove J, Leppälä K, et al. (2019): Genome-wide association study implicates CHRNA2 in cannabis use disorder. Nat Neurosci. 22:1066–1074.

38. Johnson EC, Demontis D, Thorgeirsson TE, Walters RK, Polimanti R, Hatoum AS, et al. (2020): A large-scale genome-wide association study meta-analysis of cannabis use disorder. The Lancet Psychiatry. 7:1032–1045.

39. Cheng Z, Zhou H, Sherva R, Farrer LA, Kranzler HR, Gelernter J (2018): Genome-wide Association Study Identifies a Regulatory Variant of RGMA Associated With Opioid Dependence in European Americans. Biol Psychiatry. 84:762–770.

40. Gelernter J, Kranzler HR, Sherva R, Koesterer R, Almasy L, Zhao H, et al. (2014): Genome-Wide Association Study of Opioid Dependence: Multiple Associations Mapped to Calcium and Potassium Pathways. Biological Psychiatry. 76:66–74.

41. Polimanti R, Walters RK, Johnson EC, McClintick JN, Adkins AE, Adkins DE, et al. (2020): Leveraging genome-wide data to investigate differences between opioid use vs. opioid dependence in 41,176 individuals from the Psychiatric Genomics Consortium. Mol Psychiatry. 25:1673–1687.

42. Deak JD, Zhou H, Galimberti M, Levey DF, Wendt FR, Sanchez-Roige S, et al. (2022): Genome-wide association study in individuals of European and African ancestry and multi-trait analysis of opioid use disorder identifies 19 independent genome-wide significant risk loci. Molecular Psychiatry. 27:3970–3979.

43. Kember RL, Vickers-Smith R, Xu H, Toikumo S, Niarchou M, Zhou H, et al. (2022): Cross-ancestry meta-analysis of opioid use disorder uncovers novel loci with predominant effects in brain regions associated with addiction. Nature Neuroscience. 25:1279–1287.

44. Sey NYA, Hu B, Iskhakova M, Lee S, Sun H, Shokrian N, et al. (2022): Chromatin architecture in addiction circuitry identifies risk genes and potential biological mechanisms underlying cigarette smoking and alcohol use traits. Mol Psychiatry. 27:3085–3094.

45. Finucane HK, Reshef YA, Anttila V, Slowikowski K, Gusev A, Byrnes A, et al. (2018): Heritability enrichment of specifically expressed genes identifies disease-relevant tissues and cell types. Nature Genetics. 50:621–629.

46. Klausen MK, Jensen ME, Møller M, Le Dous N, Jensen A, Zeeman VA, et al. (2022): Exenatide once weekly for alcohol use disorder investigated in a randomized, placebo-controlled clinical trial. JCI Insight. 7.

47. Wium-Andersen IK, Wium-Andersen MK, Fink-Jensen A, Rungby J, Jørgensen MB, Osler M (2022): Use of GLP-1 receptor agonists and subsequent risk of alcohol-related events. A nationwide register-based cohort and self-controlled case series study. Basic Clin Pharmacol Toxicol. 131:372–379.

48. Hendershot CS, Bremmer MP, Paladino MB, Kostantinis G, Gilmore TA, Sullivan NR, et al. (2025): Once-Weekly Semaglutide in Adults With Alcohol Use Disorder: A Randomized Clinical Trial. JAMA Psychiatry. 82:395–405.

49. Finucane HK, Bulik-Sullivan B, Gusev A, Trynka G, Reshef Y, Loh P-R, et al. (2015): Partitioning heritability by functional annotation using genome-wide association summary statistics. Nature Genetics. 47:1228–1235.

50. Su C, Gao L, May CL, Pippin JA, Boehm K, Lee M, et al. (2022): 3D chromatin maps of the human pancreas reveal lineage-specific regulatory architecture of T2D risk. Cell Metabolism. 34.

51. Patel H, Espinosa-Carrasco J, Langer B, Ewels P, Bot N-C, Garcia MU, et al. (2023): nf-core/atacseq: [2.1.2] - 2022-08-07. 2.1.2 ed: Zenodo.

52. Zhang H, Song L, Wang X, Cheng H, Wang C, Meyer CA, et al. (2021): Fast alignment and preprocessing of chromatin profiles with Chromap. Nature Communications. 12:6566.

53. Wingett S, Ewels P, Furlan-Magaril M, Nagano T, Schoenfelder S, Fraser P, et al. (2015): HiCUP: pipeline for mapping and processing Hi-C data. F1000Res. 4:1310.

54. Cairns J, Freire-Pritchett P, Wingett SW, Várnai C, Dimond A, Plagnol V, et al. (2016): CHiCAGO: robust detection of DNA looping interactions in Capture Hi-C data. Genome Biology. 17:1–17.

55. Open2C, Abdennur N, Fudenberg G, Flyamer IM, Galitsyna AA, Goloborodko A, et al. (2024): Pairtools: From sequencing data to chromosome contacts. PLOS Computational Biology. 20:e1012164.

56. Abdennur N, Goloborodko A, Imakaev M, Kerpedjiev P, Fudenberg G, Oullette S, et al. cooler.

57. Imakaev M, Fudenberg G, McCord RP, Naumova N, Goloborodko A, Lajoie BR, et al. (2012): Iterative correction of Hi-C data reveals hallmarks of chromosome organization. Nature Methods. 9:999–1003.

58. Roayaei Ardakany A, Gezer HT, Lonardi S, Ay F (2020): Mustache: multi-scale detection of chromatin loops from Hi-C and Micro-C maps using scale-space representation. Genome Biology. 21:1–17.

59. A K, S B, F A (2020): Identifying statistically significant chromatin contacts from Hi-C data with FitHiC2. Nature protocols. 15.

60. Love MI, Huber W, Anders S (2014): Moderated estimation of fold change and dispersion for RNA-seq data with DESeq2. Genome Biology. 15:550.

61. Zhu A, Ibrahim JG, Love MI (2018): Heavy-tailed prior distributions for sequence count data: removing the noise and preserving large differences. Bioinformatics. 35:2084–2092.

62. Gazal S, Finucane HK, Furlotte NA, Loh PR, Palamara PF, Liu X, et al. (2017): Linkage disequilibrium-dependent architecture of human complex traits shows action of negative selection. Nat Genet. 49:1421–1427.

63. Bulik-Sullivan B, Finucane HK, Anttila V, Gusev A, Day FR, Loh P-R, et al. (2015): An atlas of genetic correlations across human diseases and traits. Nature Genetics. 47:1236–1241.

64. Werme J, van der Sluis S, Posthuma D, de Leeuw CA (2022): An integrated framework for local genetic correlation analysis. Nature Genetics. 54:274–282.

65. Hemani G, Zheng J, Elsworth B, Wade KH, Haberland V, Baird D, et al. (2018): The MR-Base platform supports systematic causal inference across the human phenome. Elife. 7.

66. Schmidt AF, Finan C, Gordillo-Marañón M, Asselbergs FW, Freitag DF, Patel RS, et al. (2020): Genetic drug target validation using Mendelian randomisation. Nature Communications. 11:3255.

67. Rohart F, Gautier B, Singh A, Lê Cao K-A (2017): mixOmics: An R package for ‘omics feature selection and multiple data integration. PLOS Computational Biology. 13:e1005752.

68. Sey NYA, Hu B, Mah W, Fauni H, McAfee JC, Rajarajan P, et al. (2020): A computational tool (H-MAGMA) for improved prediction of brain-disorder risk genes by incorporating brain chromatin interaction profiles. Nature Neuroscience. 23:583–593.

69. Street K, Risso D, Fletcher RB, Das D, Ngai J, Yosef N, et al. (2018): Slingshot: cell lineage and pseudotime inference for single-cell transcriptomics. BMC Genomics. 19:477.

70. Littleton SH, Trang KB, Volpe CM, Cook K, DeBruyne N, Maguire JA, et al. (2024): Variant-to-function analysis of the childhood obesity chr12q13 locus implicates rs7132908 as a causal variant within the 3’ UTR of FAIM2. Cell Genom. 4:100556.

71. Koob GF, Volkow ND (2010): Neurocircuitry of Addiction. Neuropsychopharmacology. 35:217–238.

72. Koob GF, Volkow ND (2016): Neurobiology of addiction: a neurocircuitry analysis. Lancet Psychiatry. 3:760–773.

73. Koob GF (2020): Neurobiology of Opioid Addiction: Opponent Process, Hyperkatifeia, and Negative Reinforcement. Biol Psychiatry. 87:44–53.

74. Xu C, Loh HH, Law PY (2016): Effects of addictive drugs on adult neural stem/progenitor cells. Cell Mol Life Sci. 73:327–348.

75. Robinson TE, Kolb B (2004): Structural plasticity associated with exposure to drugs of abuse. Neuropharmacology. 47 Suppl 1:33–46.

76. Montoya-Filardi A, Mazón M (2017): The addicted brain: imaging neurological complications of recreational drug abuse. Radiologia. 59:17–30.

77. Bhattacherjee A, Djekidel MN, Chen R, Chen W, Tuesta LM, Zhang Y (2019): Cell type-specific transcriptional programs in mouse prefrontal cortex during adolescence and addiction. Nature Communications. 10:4169.

78. Chesi A, Wagley Y, Johnson ME, Manduchi E, Su C, Lu S, et al. (2019): Genome-scale Capture C promoter interactions implicate effector genes at GWAS loci for bone mineral density. Nature Communications. 10:1–11.

79. Su C, Argenziano M, Lu S, Pippin JA, Pahl MC, Leonard ME, et al. (2021): 3D promoter architecture re-organization during iPSC-derived neuronal cell differentiation implicates target genes for neurodevelopmental disorders. Progress in Neurobiology. 201:102000.

80. Lasconi C, Pahl MC, Pippin JA, Su C, Johnson ME, Chesi A, et al. (2022): Variant-to-gene-mapping analyses reveal a role for pancreatic islet cells in conferring genetic susceptibility to sleep-related traits. Sleep. 45.

81. Pahl MC, Doege CA, Hodge KM, Littleton SH, Leonard ME, Lu S, et al. (2021): Cis-regulatory architecture of human ESC-derived hypothalamic neuron differentiation aids in variant-to-gene mapping of relevant complex traits. Nature Communications. 12:1–12.

82. Su C, Johnson ME, Torres A, Thomas RM, Manduchi E, Sharma P, et al. (2020): Mapping effector genes at lupus GWAS loci using promoter Capture-C in follicular helper T cells. Nature Communications. 11:3294.

83. Trang KB, Pahl MC, Pippin JA, Su C, Littleton SH, Sharma P, et al. (2025): 3D genomic features across >50 diverse cell types reveal insights into the genomic architecture of childhood obesity. Elife. 13.

84. Conery M, Pippin JA, Wagley Y, Trang K, Pahl MC, Villani DA, et al. (2025): GWAS-informed data integration and non-coding CRISPRi screen illuminate genetic etiology of bone mineral density. Genome Biol. 26:331.

85. Arntfield ME, van der Kooy D (2011): β-Cell evolution: How the pancreas borrowed from the brain: The shared toolbox of genes expressed by neural and pancreatic endocrine cells may reflect their evolutionary relationship. BioEssays: news and reviews in molecular, cellular and developmental biology. 33:582–587.

86. van Arensbergen J, García-Hurtado J, Moran I, Maestro MA, Xu X, Van de Casteele M, et al. (2010): Derepression of Polycomb targets during pancreatic organogenesis allows insulin-producing beta-cells to adopt a neural gene activity program. Genome Res. 20:722–732.

87. Devaskar SU, Giddings SJ, Rajakumar PA, Carnaghi LR, Menon RK, Zahm DS (1994): Insulin gene expression and insulin synthesis in mammalian neuronal cells. J Biol Chem. 269:8445–8454.

88. Juan-Mateu J, Rech TH, Villate O, Lizarraga-Mollinedo E, Wendt A, Turatsinze JV, et al. (2017): Neuron-enriched RNA-binding Proteins Regulate Pancreatic Beta Cell Function and Survival. J Biol Chem. 292:3466–3480.

89. Ojo O, Wang XH, Ojo OO, Ibe J (2018): The Effects of Substance Abuse on Blood Glucose Parameters in Patients with Diabetes: A Systematic Review and Meta-Analysis. Int J Environ Res Public Health. 15.

90. Bruggeman BS, Campbell-Thompson M, Filipp SL, Gurka MJ, Atkinson MA, Schatz DA, et al. (2021): Substance Use Affects Type 1 Diabetes Pancreas Pathology: Implications for Future Studies. Front Endocrinol (Lausanne). 12:778912.

91. Reece AS, Hulse GK (2023): Sociodemographically Stratified Exploration of Pancreatic Cancer Incidence in Younger US Patients: Implication of Cannabis Exposure as a Risk Factor. Gastroenterology Insights. 14:204–235.

92. Kim JY, Lee DY, Lee YJ, Park KJ, Kim KH, Kim JW, et al. (2015): Chronic alcohol consumption potentiates the development of diabetes through pancreatic β-cell dysfunction. World J Biol Chem. 6:1–15.

93. Steiner JL, Crowell KT, Lang CH (2015): Impact of Alcohol on Glycemic Control and Insulin Action. Biomolecules. 5:2223–2246.

94. Nelson NG, Weingarten MJ, Law WX, Sangiamo DT, Liang N-C (2019): Joint and separate exposure to alcohol and Δ9-tetrahydrocannabinol produced distinct effects on glucose and insulin homeostasis in male rats. Scientific Reports. 9:12025.

95. Moustafa SR (2021): The immune-opioid axis in prediabetes: predicting prediabetes with insulin resistance by plasma interleukin-10 and endomorphin-2 to kappa-opioid receptors ratio. Diabetology & Metabolic Syndrome. 13:61.

96. Koekkoek LL, van der Gun LL, Serlie MJ, la Fleur SE (2022): The Clash of Two Epidemics: the Relationship Between Opioids and Glucose Metabolism. Current Diabetes Reports. 22:301–310.

97. Song D, Geetha HS, Jain S, Reyes JV, Jaiswal V, Nepal G, et al. (2022): Delayed presentation of cannabis induced pancreatitis. Clin Case Rep. 10:e05595.

98. Qi M, Kaddis JS, Chen K-T, Rawson J, Omori K, Chen ZB, et al. (2021): Chronic marijuana usage by human pancreas donors is associated with impaired islet function. PLOS ONE. 16:e0258434.

99. Cortes-Justo E, Garfias-Ramírez SH, Vilches-Flores A (2023): The function of the endocannabinoid system in the pancreatic islet and its implications on metabolic syndrome and diabetes. Islets. 15:1–11.

100. Duncan A, Heyer MP, Ishikawa M, Caligiuri SPB, Liu X-a, Chen Z, et al. (2019): Habenular TCF7L2 links nicotine addiction to diabetes. Nature. 574:372–377.

101. Meredith LR, Burnette EM, Grodin EN, Irwin MR, Ray LA (2021): Immune treatments for alcohol use disorder: A translational framework. *Brain*, Behavior, and Immunity. 97:349–364.

102. Crews FT, Vetreno RP (2016): Mechanisms of neuroimmune gene induction in alcoholism. Psychopharmacology. 233:1543–1557.

103. Erickson EK, Grantham EK, Warden AS, Harris RA (2019): Neuroimmune signaling in alcohol use disorder. Pharmacology Biochemistry and Behavior. 177:34–60.

104. Osterndorff-Kahanek EA, Becker HC, Lopez MF, Farris SP, Tiwari GR, Nunez YO, et al. (2015): Chronic Ethanol Exposure Produces Time-and Brain Region-Dependent Changes in Gene Coexpression Networks. PLOS ONE. 10:e0121522.

105. Fan X, Peters BA, Jacobs EJ, Gapstur SM, Purdue MP, Freedman ND, et al. (2018): Drinking alcohol is associated with variation in the human oral microbiome in a large study of American adults. Microbiome. 6:59.

106. Wang W, Wang Q, Qi X, Gurney M, Perry G, Volkow ND, et al. (2024): Associations of semaglutide with first-time diagnosis of Alzheimer’s disease in patients with type 2 diabetes: Target trial emulation using nationwide real-world data in the US. Alzheimer’s & Dementia. 20:8661–8672.

107. Xie Y, Choi T, Al-Aly Z (2025): Mapping the effectiveness and risks of GLP-1 receptor agonists. Nature Medicine. 31:951–962.

108. Merkel R, Moreno A, Zhang Y, Herman R, Ben Nathan J, Zeb S, et al. (2021): A novel approach to treating opioid use disorders: Dual agonists of glucagon-like peptide-1 receptors and neuropeptide Y2 receptors. Neuroscience & Biobehavioral Reviews. 131:1169–1179.

109. Cummings JL, Atri A, Feldman HH, Hansson O, Sano M, Knop FK, et al. (2025): evoke and evoke+: design of two large-scale, double-blind, placebo-controlled, phase 3 studies evaluating efficacy, safety, and tolerability of semaglutide in early-stage symptomatic Alzheimer’s disease. Alzheimer’s Research & Therapy. 17:14.

110. Du H, Meng X, Yao Y, Xu J (2022): The mechanism and efficacy of GLP-1 receptor agonists in the treatment of Alzheimer’s disease. Front Endocrinol (Lausanne). 13:1033479.

111. Edvardsson CE, Adermark L, Gottlieb S, Alfreji S, Emous TA, Gouda Y, et al. (2026): Tirzepatide reduces alcohol drinking and relapse-like behaviours in rodents. EBioMedicine. 124:106119.

112. Worth T (2024): Could the gut give rise to alcohol addiction? Nature.

113. García-Cabrerizo R, Cryan JF (2024): A gut (microbiome) feeling about addiction: Interactions with stress and social systems. Neurobiol Stress. 30:100629.

114. Bruschetta G, Diano S (2019): Brain-to-pancreas signalling axis links nicotine and diabetes. Nature. 574:336–337.

