## Supplementary methods and supplementary figure legends for "Shared and unique chromatin accessibility and 3D functional interactions of substance use disorder loci implicate cell types beyond the brain"

**SUPPLENTARY METHODS**

***Data and resource:***

**Table S1** lists the datasets we have used in prior studies. The original published studies provided configurations and technical details for ATAC-seq, Hi-C, and Capture-C library generation. The same applies for our datasets not previously published.

***ATAC-seq preprocessing and peak calling:***

Open chromatin regions (OCRs) were called using the Nextflow pipeline [*https://nf-co.re/atacseq*](https://nf-co.re/atacseq) ^(1)^. Reads were aligned to the GRCh38/hg38 assembly genome using Chromap ^(2)^, duplicates were removed, alignments from all replicates were pooled, and narrow peaks were called using MACS2. ScATAC-seq data from pancreatic cells was analyzed separately ^(3)^, fragments from the same cell ID groups were pooled into a single bam file for each cell type per sample, which were converted to fastq and used as input for the Nextflow pipeline.

***RNA-seq preprocessing and expression profiling:***

The detail configurations, steps, and technical details for each dataset are provided in the original studies. In brief, read fragments from fastq files were mapped to genome assembly GRCh38/hg38 using STAR, independently for each replicate and condition. We used GENCODE annotation files for feature annotation and htseq-count for raw read count calculation at each feature. Read counts were transformed into TPM (transcript per million) and normalized internally between replicates/conditions in each individual study. For comparative measurements, we transformed all the expression values into 0-100 scale.

***Promoter Capture-C pre-processing and interaction calling:***

Paired-end reads were pre-processed using the HICUP pipeline ^(4)^ with bowtie2 and GRCh38/hg38. Significant promoter interactions were called using unique read pairs using CHICAGO ^(5)^. We analyzed individual fragments (1frag) and binned four fragments to improve long-distance sensitivity^(6)^. Interactions with CHICAGO score > 5 at either 1-fragment or 4-fragment resolution were considered significant. Interactions from both resolutions were merged.

***Hi-C pre-processing and interaction calling:***

As described in our recent study ^(3)^, paired-end reads from each replicate were pre-processed using the HICUP v0.7.4 pipeline ^(4)^ and aligned by bowtie2 with GRCh38/hg38. The alignment files were parsed and processed by pairtools v0.3.0 ^(7)^ and indexed and compressed by pairix v0.3.7 ^(8)^, then converted to Hi-C matrix binary format .*cool* at multiple resolutions (500 bp; 1, 2, 4, 10, 40, and 500 kbp; and 1 Mbp) by cooler v0.8.11 ^(9)^ and normalized using the ICE method ^(10)^. To demonstrate data quality and the effective resolution range, a distance-decay plot was generated (**Figure S9**). The matrices from different replicates were merged at each resolution using cooler. Significant cis-interaction loops were called from the merged replicate matrices using both Mustache v1.0.1 ^(11)^ (*P*-value threshold < 0.1) and Fit-Hi-C2 v2.0.7 ^(12)^ (FDR < 1×10^−6^). This analysis was performed at three specific resolutions: 1 kb, 2 kb, and 4 kb. We selected this resolution range as it is commonly used for studying chromatin loops, consistent with recommendations from both tools, and provides a suitable balance for detecting both short- and long-range promoter-cRE interactions, particularly within TADs, which aligns with the focus of our study. The identified interaction loops were merged between the two tools, and then merged across resolutions, prioritizing higher resolution for overlaps.

The identification of chromatin interactions can be influenced by the choice of computational algorithms and parameters. Our primary analysis already incorporated measures to ensure robustness:

- **Hi-C Data:** We utilized two statistically distinct loop callers, **Mustache** and **Fit-Hi-C2**, at multiple resolutions (1kb, 2kb, 4kb). A high-confidence consensus loop set was generated by merging interactions identified by both methods, thereby mitigating biases associated with any single algorithm.
- **Promoter Capture-C Data:** We employed **CHICAGO**, the standard tool specifically designed for capture Hi-C data, and analyzed interactions at multiple sensitivities (1-fragment and 4-fragment bins) to capture both short- and long-range promoter contacts reliably.

This multi-algorithm and multi-resolution strategy were chosen *a priori* to enhance the reliability of the identified interactions used for downstream annotation and enrichment analyses. Therefore, a separate sensitivity analysis testing alternative callers was deemed redundant given the robustness incorporated into the primary workflow.

***Definition of Specifically Expressed Genes set (SEGs):***

Normalized transcripts per million (TPM) of all measured genes in 46 of 59 cell types was used to perform differential analysis using *DEseq2* package ^(13)^, where cell type and system (immune, metabolic, neural and other) were used as variables for the modeling contrast. we used *apeglm* method for effect size (logarithmic fold change estimates) shrinkage ^(14)^ to alleviate this phenomenon during the genes ranking. We followed the original method ^(15)^, chose 10% of the highest-ranking genes (3000-5000) for each cell type, added 100kb windows on either side of the transcribed region of each gene in the set of specifically expressed genes to construct a “SEGs” annotation corresponding to that cell type.

***Reformatting of the GWAS summary statistics:***

**Table S2** lists the studies from which we drew European-population GWAS summary statistics for each of the four SUDs and all other traits and disorders we explored in this study. We applied *--merge-alleles* with the list of HapMap3 variants to standardize all the GWAS summary statistics files. The baseline model LD scores, plink files, allele frequencies, HapMap3 variants list and regression weight files for the European 1000 genomes project phase 3 in GRCh38 <https://alkesgroup.broadinstitute.org/LDSCORE/GRCh38/>.

***Cell type specific partitioned heritability of each trait****:*

We used LDSC v.1.0.1 with *--h2* flag ^(16)^ to estimate the SNP-based heritability of each trait. We categorized the genome based on chromatin accessibility and interactions from each cell type into:

- **Total OCR:** For a global assessment of enrichment.
- **Promoter OCR:** To test the hypothesis that GWAS variants may impact gene expression by directly affecting promoter activity.
- **cREs, and cREs ± 500bp:** To evaluate the contribution of distal regulatory elements, such as enhancers and suppressors identified by chromatin loops, to GWAS.
- **"not-cREs/Prom OCRs":** As a negative control to ensure the specificity of enrichment in regulatory regions, we also examined **open chromatin regions not locate at promoters or having chromatin loop**, where we expected to observe minimal signal.
- **SEGs**: as positive controls, sets of cell-type-specifically expressed genes^(15)^ for each cell type.

Each set of input regions from each cell type was used to create the annotation, which in turn was used to compute annotation-specific LD scores for each cell type region of interest. To statistically account for potential confounding due to physical overlaps between our annotations and other functional genomic elements, these annotation-specific LD scores were analyzed jointly with the 63 functional categories included in the full baseline-LD model (v2.2). This comprehensive baseline model, incorporating annotations for features like promoters, enhancers, and conserved regions, allows the S-LDSC regression framework to partition heritability appropriately, attributing shared variance and mitigating potential inflation of enrichment estimates due to annotation overlap. Two statistics we used to assess how effectively our annotations capture causal variation are partitioned heritability enrichment and standardized effect size (**τ***), as previously defined^(17)^. Partitioned heritability enrichment (with its associated *P*-value) is a statistical test used to determine if a category of genomic elements is significantly associated with SUD heritability. The standardized effect size (τ*) is a quantitative measure that allows for estimating the magnitude of this contribution and enables it to be comparable across different cell types and annotation sizes. When conditioning two correlated annotations in a joint S-LDSC model, they may show similar enrichments, but the τ* for the annotation with a higher true causal variant membership will be larger and more positive.

To confirm the robustness of our enrichment findings to potential technical confounders, sensitivity analyses were performed, where we down-sampled libraries to uniform depths (see **Sensitivity analysis** for details).

Importantly, the S-LDSC model intrinsically accounts for the genomic size of each annotation category, ensuring that enrichment estimates and *P*-values are comparable across categories of different sizes. The standard S-LDSC workflow output logs confirmed all our annotation categories passed the algorithm’s default quality control checks: spanning at least 1.7% of the 0.01 centimorgan (cM) blocks of the genome (recommended minimum for one-sided enrichment tests), and at least 8.3% of the 0.01 cM blocks of the genome (recommended minimum for two-sided tests of the regression coefficient). This check verifies that each annotation is sufficiently large and distributed across the genome to support valid statistical inference and control for Type 1 error.

***Genetic correlation analysis:***

We used LDSC with --rg flag ^(18)^ to compute the global genetic correlations (r_g_) between each of the SUDs and other traits and disorders using European-ancestry meta-analysis summary statistics from the most recent GWAS (**Table S2**). The global genetic correlation between each pair of traits was computed with unconstrained intercepts, and standardized variants from the GWAS of each trait. We then partitioned the variants of each trait into cREs of each cell type and recomputed the cell type-specific genetic correlation between each SUD and other traits.

***Local genetic correlation analysis****:*

*LAVA (Local Analysis of [co]Variant Association) v0.1.0* ^(19)^ was used to estimate genetic correlations within local genomic regions across 10 pair-wise combinations of 4 SUDs and T2D. It accounts for the correlation across LD SNPs by converting marginal SNP effects into their combined effects, based on an external LD reference. The reference genome data were the same as *LDSC* analysis, however we used the GWAS without *--merge-alleles* with the list of HapMap3. For locus definition, we followed the LAVA partitioning algorithm manual ([*https://github.com/cadeleeuw/lava-partitioning*](https://github.com/cadeleeuw/lava-partitioning)) with default parameters to partition human genome build 38 into blocks of approximately equal size (~1 Mb) while minimizing the LD between them, resulting in 7351 LD blocks, excluding the extended MHC region chr6: 28,510,120-33,480,577. To account for the sample overlap, we used the pair-wise intercepts from the bivariate global LDSC to create a symmetric matrix, then converted it to a correlation matrix and provided it to LAVA. For each pair of traits, if a locus in which both traits exhibited univariate signals (genetic contribution of each single trait) passed a significance threshold of FDR<0.05, then bivariate analysis was performed to quantify the shared genetic influence on two traits. 7,343 loci were detected in univariate analysis as simultaneously significant in at least one pair of traits, and then subjected to bivariate testing, resulting in 20582 bivariate tests conducted in total (**Table S4**). A correlation between two traits at one locus was considered significant if *P*-value < 0.05/20582 (equivalent of FDR<0.05).

***sPLS-DA and DIABLO Analysis:***

To investigate the shared genetic pathways and regulatory mechanisms underlying the observed genetic overlap between T2D and SUDs, particularly focusing on pancreatic β-cells and neural cell types, we employed multivariate dimension reduction and integration techniques using the *mixOmics* ^(20)^ (v6.18.0) R package. Initial exploratory analysis of cell-type-specific gene expression profiles was performed using *sparse Partial Least Squares Discriminant Analysis (sPLS-DA)*. This supervised method was applied to RNA-seq data from six cell types: cortical neurons, neural progenitors (both iPSC-derived and ESC-derived), and pancreatic cells α and β-cells. This analysis helped to visualize the relationships between different cell types based on their transcriptomic signatures and shared expression patterns. To integrate the transcriptomic and chromatin accessibility data within the genomic loci exhibiting significant genetic correlation between T2D and SUDs (specifically TUD), we utilized the *Data Integration Analysis for Biomarker discovery using Latent cOmponents (DIABLO)* framework. This supervised integration approach was applied to the subset of genes and OCRs, including promoter OCRs and cREs, located within the identified correlated loci. These tools were built to discriminate between different groups (cell types in our case) based on their molecular profiles. They aim to find latent components that maximize the covariance between the omics data and the categorical outcome (cell types), thereby enhancing the separation of cell types.

***Cell-Type Specific H-MAGMA Analysis:***

To prioritize biological candidate genes underlying the shared genetic architecture of SUDs, we performed a chromatin-interaction-informed gene association analysis using H-MAGMA ([*https://github.com/thewonlab/H-MAGMA*](https://github.com/thewonlab/H-MAGMA))(22), allowing for the identification of regulatory risk variants located in distal enhancers. We constructed custom MAGMA annotation files for each of the cell types utilizing our cell type-specific Capture C, Hi-C, and ATAC-seq data, by intersecting GWAS variants with cell-type-specific cREs and linking them to target promoters via chromatin interactions. We performed a gene-level association analysis for each trait-cell type combination using MAGMA (v1.08). The gene-level test statistic was computed by aggregating the P-values of all SNPs mapped to a gene’s regulatory landscape (exons, promoter, and linked distal cREs), adjusting for gene size, SNP density, and local LD structure using the 1000 Genomes Phase 3 European reference panel. To ensure comparative validity across traits with varying GWAS statistical power, we applied a uniform FDR correction. Genes were considered significantly associated if they met a threshold of FDR < 0.05 within each specific analysis.

***Pathway Enrichment Analysis*:**

Following the identification of key genes and regulatory elements through sPLS-DA and DIABLO analyses, pathway enrichment analysis was performed using the *pathfinder* ^(23)^ and *clusterProfiler* ^(24)^ R package. Genes identified as significantly contributing to the separation or integration patterns in the sPLS-DA and DIABLO models were used as input. These tools leverage active subnetworks to identify enriched pathways and gene sets from various databases (e.g., KEGG, Reactome, GO), providing a functional interpretation of the molecular features highlighted by the multivariate analyses. This step aimed to consolidate the list of individual genes and regulatory elements into broader biological contexts, revealing the specific pathways and mechanisms potentially shared between T2D and SUDs in the studied cell types.

***Signaling Inference and Network Construction with CellChat*:**

To map the intercellular signaling architecture of the "Pancreatic-SUD Axis," we utilized CellChat (v2.2.0) to infer, visualize, and analyze cross-tissue ligand-receptor interactions. Using the curated CellChatDB.human repository, comprising 3,300+ validated molecular interactions across Secreted Signaling, ECM-Receptor, and Cell-Cell Contact categories, we integrated our analysis to "Secreted Signaling" pathways.

First, we identified conditionally significant risk genes for each cell type using the H-MAGMA outputs (FDR ≤ 0.05). To isolate the directed neuro-endocrine axis, we cross-referenced pancreatic risk genes (from α- and β- cells) with the CellChat database to identify putative "Sender" ligands. Independently, we cross-referenced neural risk genes (derived from hESC- and iPSC-lineage NPCs and neurons) with the database to identify corresponding "Receiver" receptors. We then mapped valid signaling interactions, retaining only pairs consisting of a pancreatic-specific risk ligand matching a neural-specific risk receptor.

To determine the statistical significance of these directed neuro-endocrine interactions, we conducted gene-set enrichment analysis using MAGMA. The identified ligand-receptor pairs were formatted as custom gene sets (--set-annot) and evaluated against the primary cell-type-specific H-MAGMA output (--gene-results), utilizing the comprehensive, unannotated H-MAGMA gene list as the background model for permutations. The resulting MAGMA pathway P-values were converted into normalized Z-scores Z = Φ^-1^(1 - *P*). For each trait (AUD, CanUD, OUD, TUD, and T2D), we calculated the mean aggregate Z-score and standard error for the overall pancreatic ligand burden and the neural receptor burden.

Pair-Specific Genetic Burden Chord Visualization: We defined impacted Interactions as specific signaling pathways where both the encoding ligand from pancreatic cells and its cognate receptor from neural cells exhibited significant genetic burden with significance defined at P-values < 0.05 within their respective cell-type-specific contexts. We extracted the gene-level Z-statistics for these specific targets from the MAGMA outputs. For each valid interaction, we calculated a combined interaction burden score by summing the pancreatic ligand Z-score and the neural receptor Z-score. The relative genetic burden of specific neuro-endocrine signaling families (INH/ACVR, SEMA/NRP/PLXN, UCN/CRHR) across the different SUDs and T2D was visualized using network chord diagrams (***circlize***)

***Developmental Trajectory Mapping and Kinetic Risk Modeling***:

To reconstruct the temporal emergence of genetic liability, we modeled trait-associated risk scores across a neural differentiation continuum. This analysis utilized a single-cell transcriptomic reference of hESC-derived hypothalamic lineages, spanning the transition from undifferentiated stem cells to mature neuronal and glial populations.

We performed trajectory analysis using Slingshot^(25)^ to root the lineage in hypothalamic stem cells. Cells were ordered along a continuous pseudotime axis representing the differentiation path through neural progenitors (NPCs) toward terminal fates. To ensure biological relevance, we cross-validated these trajectories against a pediatric hypothalamic single-cell atlas (ages 4-14) ^(26)^, specifically monitoring the maturation of hypothalamic-specific markers.

We stratified risk genes identified via H-MAGMA into distinct functional modules based on their peak expression in the reference dataset. For each disorder (TUD, AUD, CanUD, OUD), we defined three cell-type-specific modules:

- **Neuron Module:** Genes characterized by peak expression in mature neuronal clusters.
- **Progenitor Module:** Genes associated with fetal proliferative and early commitment states.
- **Astrocyte Module:** Genes specific to the macroglial lineage.

Individual cell-level **Risk Scores** were calculated by aggregating the expression of genes within these modules using the AddModuleScore() function in Seurat.

To quantify the "kinetics" of risk, we integrated the calculated scores with the pseudotime values and metadata. We modeled the dynamics of each module, such as Score_TUD_Neuron or Score_AUD_Progenitor, along the differentiation axis. This allowed us to distinguish between three distinct kinetic mechanisms:

- **Switch-like Surge:** A sharp increase in the Progenitor module during late differentiation, indicating the repurposing of developmental genes for mature function.
- **Constitutive Elevation:** Early and sustained elevation of risk scores from the stem cell stage onward.
- **Maturation-Dependent:** Risk scores that remain negligible until terminal differentiation, tied to the acquisition of mature synaptic machinery.

The relationship between pseudotime and risk scores was smoothed using General Additive Models (GAMs) to identify significant inflection points where genetic liability emerged.

***Enrichment patterns across varying open chromatin region definitions***

We assessed the cell-type specific enrichment of GWAS signals in specific genomic regions: total open chromatin regions (total OCRs), promoter-associated open chromatin regions (promoter OCRs), and putative regulatory elements contacting promoters via chromatin loops (cREs, the putative active and poised enhancers/silencers). We hypothesized that trait-associated variants are significantly enriched in cREs and promoter OCRs, reflecting their potential roles in gene regulation (e.g., altering enhancer or promoter activity). Conversely, we expected minimal enrichment in open chromatin regions not classified as cREs or promoters ("not-cREs/Prom OCRs"), which served as negative controls, while annotations based on the top 10% of cell-type-specifically expressed genes plus 100kb flanking regions (SEGs, as defined in Methods) served as positive controls expected to capture cell-type-specific signals. Consistent with the approach of the original S-LDSC method, which modeled 500-bp flanking regions separately to account for signals in adjacent areas ^(16)^, we chose to expand our core cRE annotations by ±500 bp. This expansion aimed to capture potential causal variants residing near, but not directly within, peak summits, a common occurrence in regulatory genomics. While this approach increases the number of variants considered, incorporating nearly as many weighted variants into the enrichment calculation as the broader 'total OCRs' category in most instances, we evaluated its impact empirically. For GWAS signal distributions with sharp peaks, expanding the region by 500 bp has been reported to dilute the signal and increase *P*-values and enrichment standard errors without increasing heritability enrichment^(27)^.

First, we tested whether specific categories of genomic elements were significantly enriched for SUD heritability using the partitioned heritability enrichment test and its associated *P*-value (**Figure S1**). For all cell types, the variants within the total set of OCRs (**“Total OCRs”**) showed a positive risk heritability (enrichment >1) for at least one SUD, including in 52 cell types each for AUD, TUD and CanUD, and 50 for OUD. The total set of implicated cell types across all four SUDs consisted of 9 distinct cell types (7 for TUD, 4 for AUD, 5 for CanUD and 1 for OUD) were the enrichments statistically significant (*P* ≤0.05). When limiting the analysis to just promoter OCRs, the enrichments were significantly lower for immune cell types, markedly greater for metabolic and some other cell types. This OCR category also yielded the greatest variability in enrichment (enrichment standard error) in all cell types and SUDs (**Figure S2**).

We observed a positive correlation between heritability enrichment in promoter OCRs and enrichment associated with SEGs across relevant cell types (Pearson's R=0.14). This correlation was somewhat stronger when comparing the standardized effect sizes τ* (R=0.28). Notably, this relationship was less pronounced in immune and metabolic cells compared to neural cells (**Figure S3**). This lineage-specific difference suggests that the regulatory programs captured by promoter OCRs align more closely with those defined by high cell-type-specific expression in the nervous system, potentially reflecting a greater reliance on promoter-proximal regulation for defining neural cell identity compared to immune or metabolic cell types.

Compared to the analysis of promoter OCRs, restricting the heritability enrichment assessment to cREs yielded greater enrichment for most of the cell types for all disorders, lower *P*-values, and more precise estimates (i.e., smaller standard errors) across different cell types. As anticipated, the expansion resulted in somewhat weaker enrichment estimates and broader standard errors compared to core cRE regions. However, it consistently yielded more significant *P*-values across most enriched cell types (**Figure S2**). This suggests that the larger annotation, while potentially less specific at the per-base level, captures a greater total amount of heritable signal relevant to these SUDs, leading to stronger overall evidence against the null hypothesis (i.e., lower *P*-value). Overlaps created by this expansion were statistically controlled for by the S-LDSC baseline-LD model (see Methods), which partitions heritability appropriately among correlated annotations.

Conversely, when we analyzed OCRs located outside both cREs and promoters (not-cREs/Prom OCRs) as a control assessment, we observed generally lower enrichment than for the cREs with their expanded regions. The inconsistencies observed highlight that even when considering SEGs and “not-cREs/Prom OCRs” as reference points, the other OCR categories demonstrate context-specific and metric-dependent correlations, reflecting the complex interplay between different regulatory elements and gene expression across cell types.

***Sensitivity analyses - Robustness to Sequencing Depth Variation:***

To assess the robustness of our main findings to potential technical confounders, we performed several sensitivity analyses. Differences in sequencing depth between Hi-C, Promoter Capture-C (PCC), and ATAC-seq libraries across different cell types could potentially influence interaction/peak calling and subsequent heritability enrichment estimates. To address this concern, we performed a down-sampling sensitivity analysis.

- **Cell Type Selection:** We selected a representative subset of cell types exhibiting high, moderate, and low/no significant SUD heritability enrichment in the primary analysis.
- **Replicate Handling:** For cell types with more than two biological replicates, we first selected the two replicates with the highest sequencing depth to retain maximal signal quality while standardizing the number of replicates to n=2 across the subset.
- **Target Depth Calculation:** We determined the target sequencing depth separately for each assay type (ATAC-seq, Hi-C, PCC), using the lowest reads count.
- **Down-sampling:** Raw FASTQ files for the chosen replicates from deeper libraries were down-sampled to the calculated target depth using seqtk sample v1.5-r133, employing identical random seeds (-s100) for paired-end files to maintain read pairing.
- **Re-analysis:** The complete data processing pipeline was re-run on these uniformly down-sampled FASTQ files. This included alignment, peak/interaction calling (using the same parameters as the main analysis), annotation generation, and partitioned heritability estimation using S-LDSC with the baseline-LD v2.2 model.

The results of the down-sampling analysis are presented in **Figure S4**. As anticipated, the reduction in data led to decreased statistical power, resulting in generally larger standard errors and less significant p-values for enrichment estimates compared to the original analysis. However, the relative enrichment patterns across the tested cell types and annotation categories remained highly consistent. Notably, the point estimates for enrichment were often stronger in the down-sampled data. This suggests that SUD heritability is particularly concentrated within the highest-confidence regulatory elements that persist even at lower sequencing depths. Overall, these findings indicate that our primary conclusions regarding cell-type-specific enrichment are robust and not primarily driven by technical variations in library size.

While this reduction in data led to an expected decrease in statistical power (i.e., less significant *P*-values), the relative enrichment patterns across cell types remained consistent, and the enrichment estimates themselves were often stronger in the down-sampled data (**Figure S4**). These results indicate that the observed cell-type-specific enrichments are robust biological signals, not primarily driven by technical differences in library size.

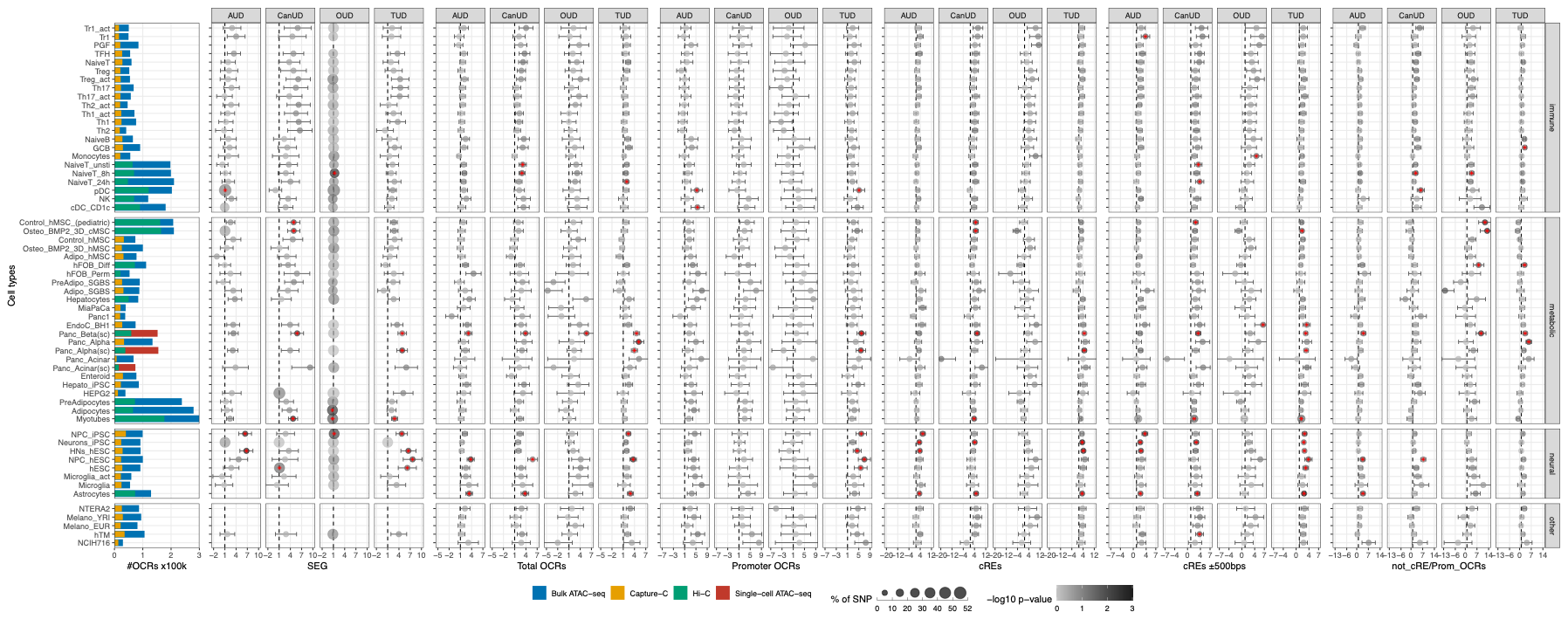

Figure S1. **S-LDSC enrichment for SUDs across diverse cell types across all annotation**s**.** Bar plots (left) is the same in Figure 1A. The dot plots depict heritability enrichment for each cell type across 4 SUD traits as determined by LDSC analysis. Whiskers represent enrichment standard errors, with colors matched for Hi-C vs. Capture-C. The colors of the dots correspond to *P* -values in -log10 of each disorder, with dots featuring a red asterisk indicating a significant FDR ≤ 0.05. The size of the dots corresponds to the proportion of SNP contribution to heritability. Dashed line at 1 indicates no enrichment.

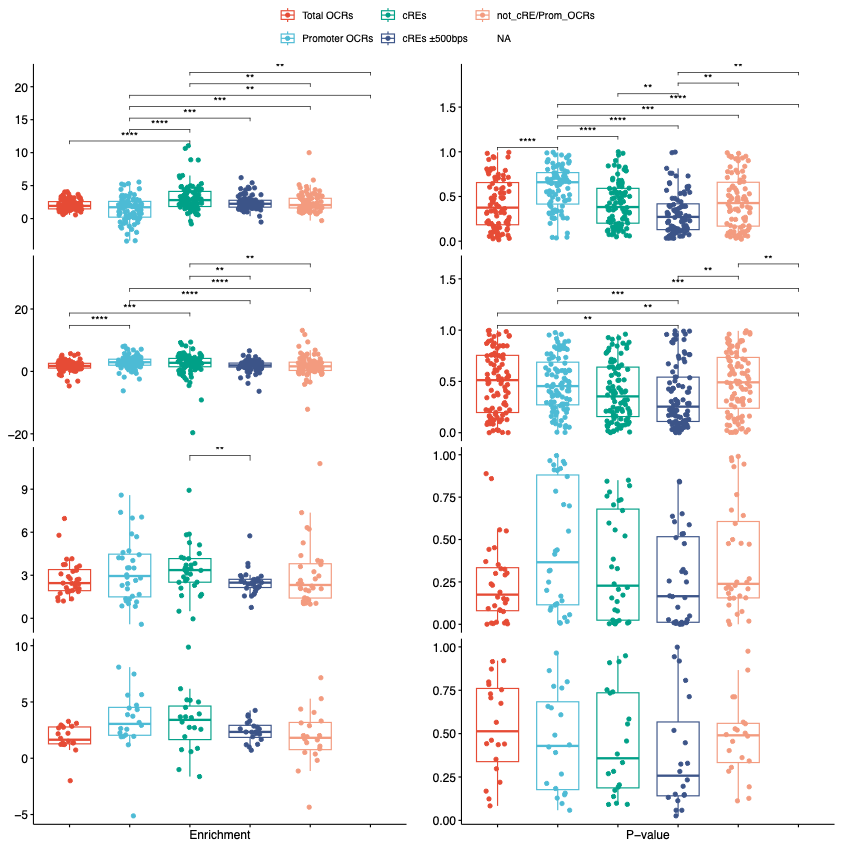

Figure S2. **Distributions of different metrics from S-LDSC** within each cell type system, colored by annotations, with two-tailed T-test *P*-values for each pairwise comparison: ****, <0.001, ***, <0.005, **, <0.01 and * <0.05

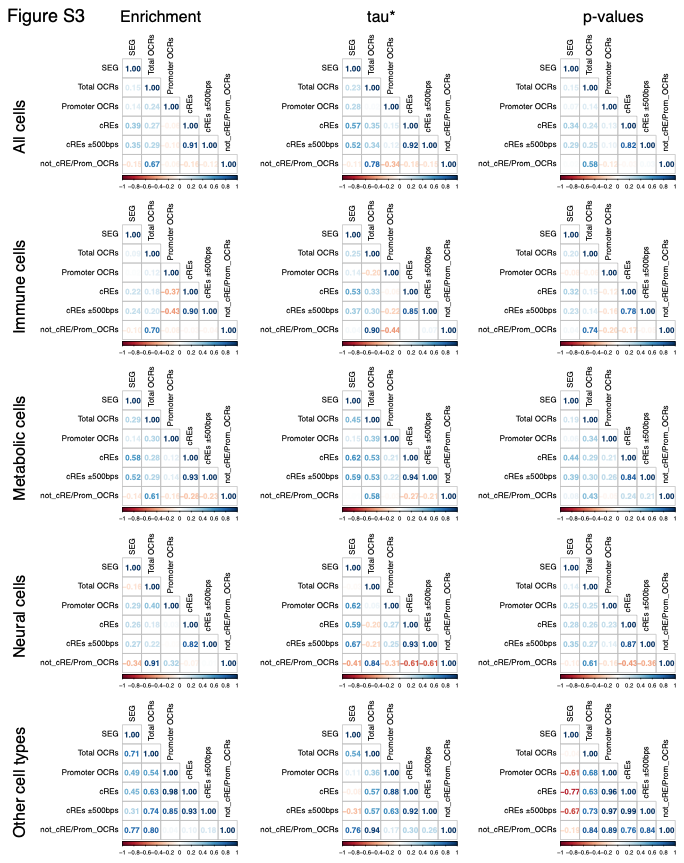

Figure S3. **Correlation between pair-wise annotations** for 3 S-LDSC metrics: enrichment, τ*, and *P*-values, across all cell types and segregated into cell systems. The correlation between SEG and not_cRE/Prom_OCRs in neural cells is strongly negative using τ* (r^2^=-0.41) aligning with expectations, yet it is positive in “other cell types” (r^2^=0.76), which is counterintuitive when comparing positive and negative control category, suggesting that distinct additional functions of the GWAS variants attributed to the phenotype are present in different cellular system settings.

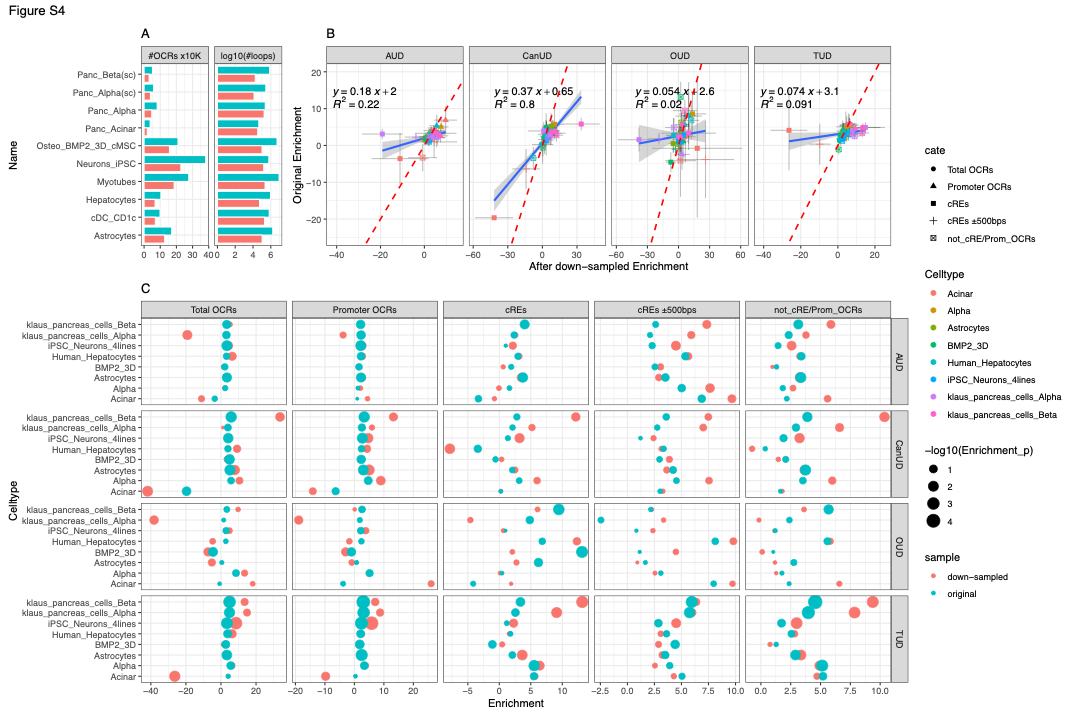

Figure S4. **Comparison of S-LDSC enrichment estimates before and after library down-sampling**. **A.** Bar-plots show number of OCRs and number of chromatin loops before and after down-sampling sequencing libraries (number of loops were scaled to log10 for easy view). **B.** This scatter plot compares the S-LDSC partitioned heritability enrichment estimates for key annotation categories before (y-axis) and after (x-axis) down-sampling sequencing libraries to a uniform depth. Each point represents a specific cell type and regulatory element category, separated by 4 panels of SUDs. The dashed red line indicates y=x, representing perfect agreement between the original and down-sampled estimates. Points lying above the line indicate a stronger enrichment estimate after down-sampling. Blue lines and their formulars noted on top show real correlations between original and down-sampled estimates. The overall positive correlation demonstrates the consistency of relative enrichment patterns, supporting the robustness of the findings to sequencing depth. C. Dot-plots show Enrichment levels of each cell types, colored by original vs down-sampled, sized by -log10(*P-*values). Majority of down-sampled estimates are more significant with smaller *P*-values.

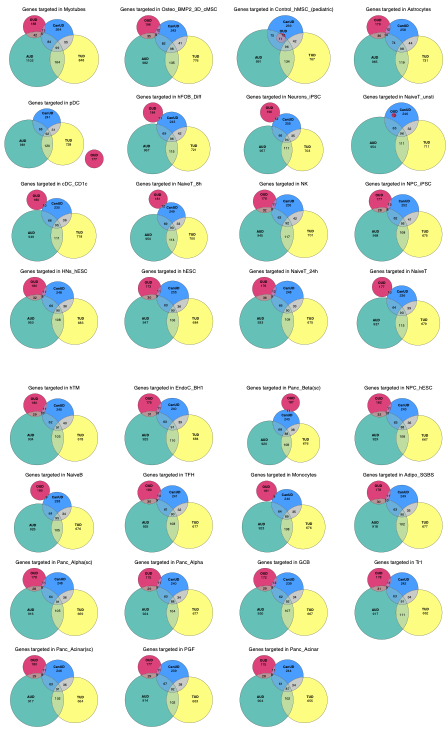

Figure S5. Venn Diagrams of risk genes identified by H-MAGMA for each SUD, performed on each of 31 cell types that showed significantly conditional effect size (τ*) in at least one annotation category.

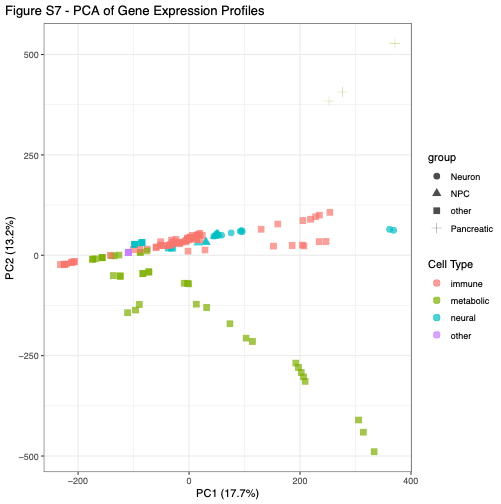

Figure S6. **Principal Component Analysis of gene expression profiles across all 59 cell types**, performed on the normalized expression values. Each point represents an individual cell type. PC1 and PC2 represent the first two principal components, capturing the largest sources of variance in the gene expression data. The six key cell types implicated in the Pancreatic-SUD axis (iPSC-derived cortical neurons, neural progenitors, ESC-derived hypothalamic NPCs, neurons, pancreatic alpha cells, and pancreatic beta cells) are highlighted as different shapes.

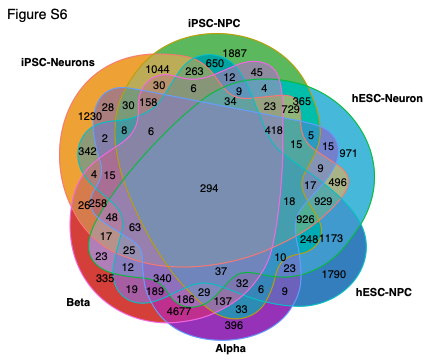

Figure S7. **Venn diagram of top 10% specially expressed genes** from 4 neural cell types and 2 pancreatic cell types, numbers are genes within each corresponding intersection.

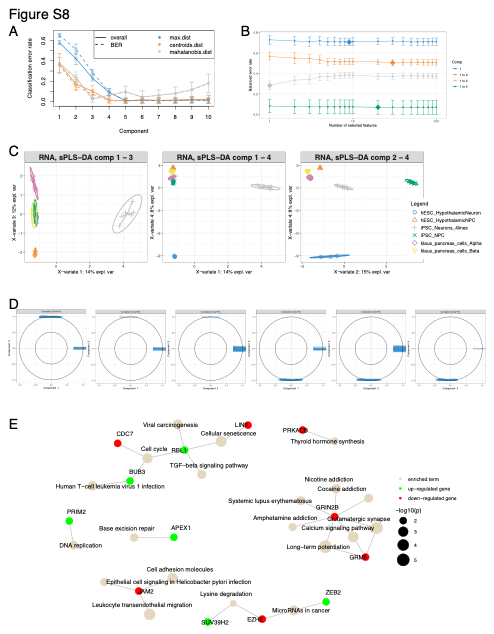

Figure S8. **Sparse Partial Least Squares discriminant analysis (sPLS-DA) on whole expression profiles of 4 neural and 2 pancreatic cell types. A.** Optimizing the number of components in sPLS-DA for the gene expression data**.** For each component, repeated 50 iterations x3-fold cross-validation (leave-one-out) is used to evaluate the PLS-DA classification performance (overall and balanced error rate BER), for each type of prediction distance  (max.dist, centroids.dist and mahalanobis.dist); bars show the standard deviation across the repeated folds; also shows that the error rate reaches a minimum from 4 components. **B.** Tuning keepX for the sPLS-DA performed on the gene expression data, each colored line represents the balanced error rate (y-axis) per component across all tested keepX values (x-axis) with the standard deviation based on the repeated cross-validation folds. The diamond indicates the optimal keepX value on a particular component which achieves the lowest classification error rate as determined with a one-sided T-test, values represented for a given component (e.g. comp 1 to 2) include the optimal keepX value chosen for the previous component (comp 1); optimal keepX parameter according to minimal error rate are 9 (comp1), 30 (comp2), 1 (comp3) and 20 (comp4). **C**. Samples are projected into the space spanned by the first four components, showing the rest of the combinations from Fig. 3A, none of the combinations of all 4 components enables us to discriminate all cell types. **D.** Correlation circle plot representing the genes selected by sPLS-DA, truncated to the first 10 characters, for each combination of pair-wise components; the plots show complete polarized loadings where the nominated genes drive the discrimination force for singular component. **E.** Network of enriched terms and genes from Fig 3B., up/down correspond to direction of fold-change expression of the discriminated cell types versus the rest.

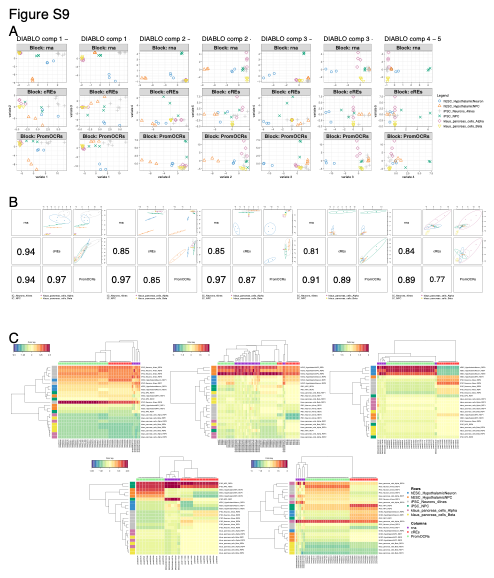

Figure S9. **Data Integration Analysis for Biomarker discovery using Latent cOmponents (DIABLO) framework using 3 blocks of RNA-seq profiles and ATAC-seq profiles from Promoter OCRs and cREs within 114 loci displayed significant correlation between T2D and SUDs. A.** Sample plot from multiblock DIABLO analysis showing the remaining combinations of 5 optimized components, complemented for Fig 3C. **B.**  Diagnostic plot where cell types are represented based on the specified component for each data set (mRNA, Promoter OCRs and cREs), bottom triangle numbers indicate the correlation coefficients between the block data sets, cREs correlated best with RNA and Promoter OCRs in the first component, while Promoter OCRs and RNA correlated best in the 2^nd^ and 3^rd^ components. **C.** Clustered Image Map for the variables selected by multiblock DIABLO on each component, Euclidean distance and Complete linkage methods are used, the CIM represents samples in rows and selected features in columns (indicated by their data type at the top of the plot).

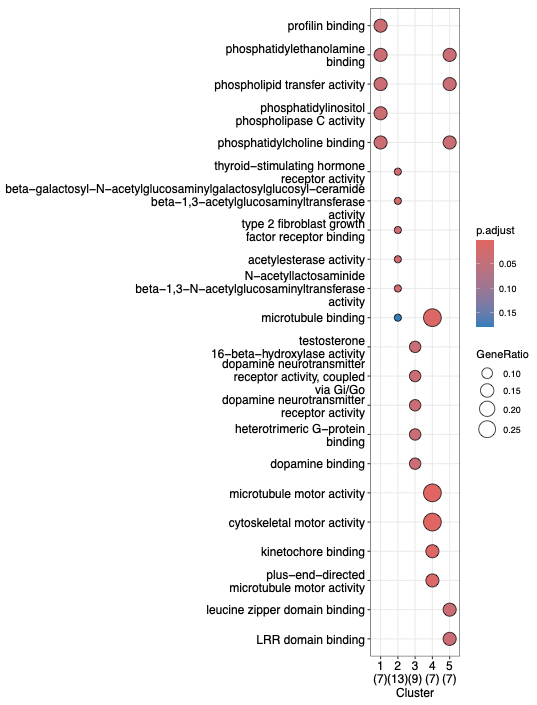

Figure S10. GO-terms enrichments of genes drove the separations with 5 optimized DIABLO components

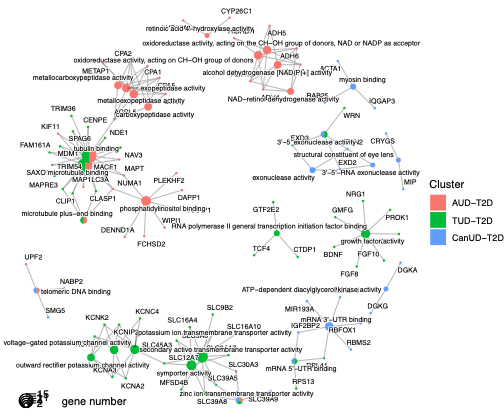

Figure S11. Network of enriched GO terms with genes from 114 loci, separated by pair-wise correlation between T2D and SUDs.

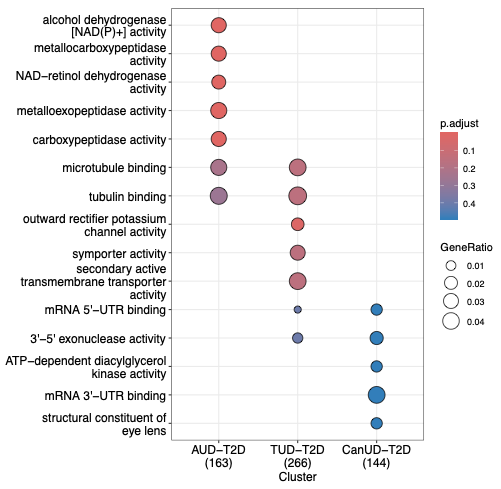

Figure S12. Enriched GO terms with genes from 114 loci, separated by pair-wise correlation between T2D and SUDs.

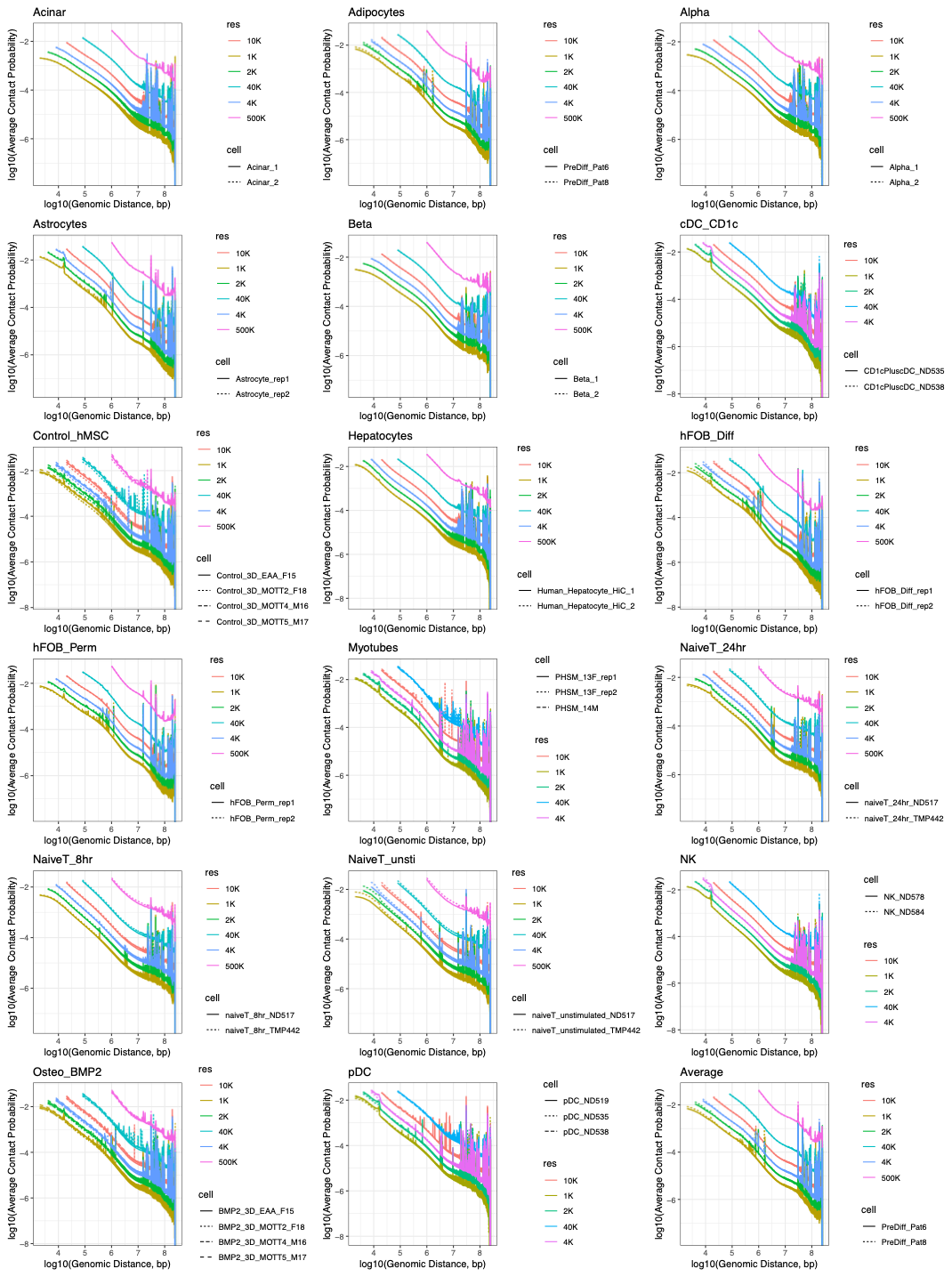

Figure S13. **Hi-C data Quality assessment via Distance-Decay plots**: Each individual panel shows the distance-decay plot for one specific cell type. The x-axis represents the linear genomic distance between bins (in base pairs, log scale), and the y-axis represents the normalized contact probability (log scale). Within each panel, each distinct line represents the distance-decay curve calculated from an individual biological replicate for that cell type. The characteristic steep downward slope observed across replicates indicates successful enrichment of cis-contacts and good quality Hi-C data.

**TABLE LEGENDS**

Table S1. **Data resources of 59 cell types**
Table S2. **4 substance use disorders and other psychiatric traits GWAS information**
Table S3. **S-LDSC results**
Table S4. **Implicated genes from H-MAGMA**
Table S5. **LAVA results**
Table S6. **MR results and diagnostics**

**References**

1.	Patel H, Espinosa-Carrasco J, Langer B, Ewels P, Bot N-C, Garcia MU, et al. (2023): nf-core/atacseq: [2.1.2] - 2022-08-07. 2.1.2 ed: Zenodo.

2.	Zhang H, Song L, Wang X, Cheng H, Wang C, Meyer CA, et al. (2021): Fast alignment and preprocessing of chromatin profiles with Chromap. *Nature Communications*. 12:6566.

3.	Su C, Gao L, May CL, Pippin JA, Boehm K, Lee M, et al. (2022): 3D chromatin maps of the human pancreas reveal lineage-specific regulatory architecture of T2D risk. *Cell Metabolism*. 34.

4.	Wingett S, Ewels P, Furlan-Magaril M, Nagano T, Schoenfelder S, Fraser P, et al. (2015): HiCUP: pipeline for mapping and processing Hi-C data. *F1000Res*. 4:1310.

5.	Cairns J, Freire-Pritchett P, Wingett SW, Várnai C, Dimond A, Plagnol V, et al. (2016): CHiCAGO: robust detection of DNA looping interactions in Capture Hi-C data. *Genome Biology*. 17:1-17.

6.	C S, MC P, SFA G, AD W (2021): Restriction enzyme selection dictates detection range sensitivity in chromatin conformation capture-based variant-to-gene mapping approaches. *Human genetics*. 140.

7.	Open2C, Abdennur N, Fudenberg G, Flyamer IM, Galitsyna AA, Goloborodko A, et al. (2024): Pairtools: From sequencing data to chromosome contacts. *PLOS Computational Biology*. 20:e1012164.

8.	S L (2020): A tool for indexing and querying on a block-compressed text file containing pairs of genomic coordinates.

9.	Abdennur N, Goloborodko A, Imakaev M, Kerpedjiev P, Fudenberg G, Oullette S, et al. cooler.

10.	Imakaev M, Fudenberg G, McCord RP, Naumova N, Goloborodko A, Lajoie BR, et al. (2012): Iterative correction of Hi-C data reveals hallmarks of chromosome organization. *Nature Methods*. 9:999-1003.

11.	Roayaei Ardakany A, Gezer HT, Lonardi S, Ay F (2020): Mustache: multi-scale detection of chromatin loops from Hi-C and Micro-C maps using scale-space representation. *Genome Biology*. 21:1-17.

12.	A K, S B, F A (2020): Identifying statistically significant chromatin contacts from Hi-C data with FitHiC2. *Nature protocols*. 15.

13.	Love MI, Huber W, Anders S (2014): Moderated estimation of fold change and dispersion for RNA-seq data with DESeq2. *Genome Biology*. 15:550.

14.	Zhu A, Ibrahim JG, Love MI (2018): Heavy-tailed prior distributions for sequence count data: removing the noise and preserving large differences. *Bioinformatics*. 35:2084-2092.

15.	Finucane HK, Reshef YA, Anttila V, Slowikowski K, Gusev A, Byrnes A, et al. (2018): Heritability enrichment of specifically expressed genes identifies disease-relevant tissues and cell types. *Nature Genetics*. 50:621-629.

16.	Finucane HK, Bulik-Sullivan B, Gusev A, Trynka G, Reshef Y, Loh P-R, et al. (2015): Partitioning heritability by functional annotation using genome-wide association summary statistics. *Nature Genetics*. 47:1228-1235.

17.	Gazal S, Finucane HK, Furlotte NA, Loh P-R, Palamara PF, Liu X, et al. (2017): Linkage disequilibrium–dependent architecture of human complex traits shows action of negative selection. *Nature Genetics*. 49:1421-1427.

18.	Bulik-Sullivan B, Finucane HK, Anttila V, Gusev A, Day FR, Loh P-R, et al. (2015): An atlas of genetic correlations across human diseases and traits. *Nature Genetics*. 47:1236-1241.

19.	Werme J, van der Sluis S, Posthuma D, de Leeuw CA (2022): An integrated framework for local genetic correlation analysis. *Nature Genetics*. 54:274-282.

20.	Rohart F, Gautier B, Singh A, Lê Cao K-A (2017): mixOmics: An R package for ‘omics feature selection and multiple data integration. *PLOS Computational Biology*. 13:e1005752.

21.	Hemani G, Zheng J, Elsworth B, Wade KH, Haberland V, Baird D, et al. (2018): The MR-Base platform supports systematic causal inference across the human phenome. *Elife*. 7.

22.	Sey NYA, Hu B, Mah W, Fauni H, McAfee JC, Rajarajan P, et al. (2020): A computational tool (H-MAGMA) for improved prediction of brain-disorder risk genes by incorporating brain chromatin interaction profiles. *Nature Neuroscience*. 23:583-593.

23.	Ulgen E, Ozisik O, Sezerman OU (2019): pathfindR: An R Package for Comprehensive Identification of Enriched Pathways in Omics Data Through Active Subnetworks. *Frontiers in Genetics*. 10.

24.	Wu T, Hu E, Xu S, Chen M, Guo P, Dai Z, et al. (2021): clusterProfiler 4.0: A universal enrichment tool for interpreting omics data. *The Innovation*. 2:100141.

25.	Street K, Risso D, Fletcher RB, Das D, Ngai J, Yosef N, et al. (2018): Slingshot: cell lineage and pseudotime inference for single-cell transcriptomics. *BMC Genomics*. 19:477.

26.	Littleton SH, Trang KB, Volpe CM, Cook K, DeBruyne N, Maguire JA, et al. (2024): Variant-to-function analysis of the childhood obesity chr12q13 locus implicates rs7132908 as a causal variant within the 3' UTR of FAIM2. *Cell Genom*. 4:100556.

27.	Trang KB, Pahl MC, Pippin JA, Su C, Littleton SH, Sharma P, et al. (2025): 3D genomic features across >50 diverse cell types reveal insights into the genomic architecture of childhood obesity. *Elife*. 13.
